# Influence of age, sex, body habitus, vaccine type and anti-S serostatus on cellular and humoral responses to SARS-CoV-2 vaccination

**DOI:** 10.1101/2023.09.29.23296222

**Authors:** Emma S Chambers, Weigang Cai, Giulia Vivaldi, David A Jolliffe, Natalia Perdek, Wenhao Li, Sian E Faustini, Joseph M. Gibbons, Corinna Pade, Alex G. Richter, Anna K Cousens, Adrian R Martineau

**Affiliations:** Centre for Immunobiology, Blizard Institute, Barts and the London School of Medicine and Dentistry, Queen Mary University of London, London E1 2AT, U.K; Institute of Immunology and Immunotherapy, College of Medical and Dental Sciences, University of Birmingham, Birmingham B15 2TT, U.K; Centre for Genomics and Child Health, Blizard Institute, Barts and the London School of Medicine and Dentristry, Queen Mary University of London, London, E1 2AT, U.K; Infectious Diseases and Immune Defence Division, Walter and Eliza Hall Institute of Medical Research, Parkville 3052, Australia; Centre for Infectious Diseases Research in Africa, Institute of Infectious Disease and Molecular Medicine, University of Cape Town, Cape Town 7925, South Africa

## Abstract

Vaccine development targeting SARS-CoV-2 in 2020 was of critical importance in reducing COVID-19 severity and mortality. In the U.K. during the initial roll-out most individuals either received two doses of Pfizer COVID-19 vaccine (BNT162b2) or the adenovirus-based vaccine from Oxford/AstraZeneca (ChAdOx1-nCoV-19). There are conflicting data as to the impact of age, sex and body habitus on cellular and humoral responses to vaccination, and most studies in this area have focused on determinants of mRNA vaccine immunogenicity. Here we studied a cohort of participants in a population-based longitudinal study (COVIDENCE UK) to determine the influence of age, sex, body mass index (BMI) and pre-vaccination anti-Spike (anti-S) antibody status on vaccine-induced humoral and cellular immune responses to two doses of BNT162b2 or ChAdOx-n-CoV-19 vaccination.

Younger age and pre-vaccination anti-S seropositivity were both associated with stronger antibody responses to vaccination. BNT162b2 generated higher neutralising and anti-S antibody titres to vaccination than ChAdOx1-nCoV-19, but cellular responses to the two vaccines were no different. Irrespective of vaccine type, increasing age was also associated with decreased frequency of cytokine double-positive CD4+ T cells. Increasing BMI was associated with reduced frequency of SARS-CoV-2-specific TNF+ CD8% T cells for both vaccines.

Together, our findings demonstrate that increasing age and BMI associate with attenuated cellular and humoral responses to SARS-CoV-2 vaccination. Whilst both vaccines induced T cell responses, BNT162b2 induced significantly elevated humoral immune response as compared to ChAdOx-n-CoV-19.

## 1. Introduction

SARS-CoV-2 is the causative agent of coronavirus disease 2019 (COVID-19). As of July 2023 SARS-CoV-2 has infected over 768 million people and is responsible for over 6.9 million deaths worldwide^1^, however this is likely to be an under-estimation as excess mortality due to COVID-19 in April 2022 was estimated to be 18 million people^2^. SARS-CoV- 2 also causes the debilitating illness, long-COVID, which affects around 10% of those infected^3^. Due to the profound morbidity and mortality caused by SARS-CoV-2, the development of vaccines in 2020 was of critical importance. In the U.K. three vaccines were licenced for use by the start of 2021. These include the mRNA vaccines from Pfizer (BNT162b2) and Moderna (mRNA-1273) and the adenovirus-based vaccine from AstraZeneca/Oxford (ChAdOx1-nCoV-19)^4^. In 2020-2021 during the initial roll-out of COVID- 19 vaccination in the U.K., most individuals received a primary course comprising two doses of BNT162b2 or ChAdOx1-nCoV-19^5, 6^.

Individuals who are male, older in age, or with high Body Mass Index (BMI) are at increased risk of morbidity and mortality from COVID-19^7, 8^ – therefore, studies have focussed on vaccine immunogenicity in these at risk populations. Increasing age has been associated with reduced IgG and neutralising antibody titre after two BNT162b2 vaccine doses^9, 10^. In contrast, a study assessing both BNT162b2 or ChAdOx1 immunogenicity showed no significant difference in humoral or cellular immune response after vaccination in those ≥80 years as compared to those <80^11^. The published BMI studies show conflicting results, with either no effect^10, 12^, decreased^13, 14^ or increased^15^ humoral BNT162b2 vaccine immunogenicity observed with increased BMI. Few studies have shown any influence of sex on vaccine efficacy – of those mentioned above, only one identified that females have a higher anti-Spike IgG antibody titre post-BNT162b2 vaccination^10^.

There are relatively few studies investigating the cellular and humoral immune response to both BNT162b2 and ChAdOx1-nCoV-19 in those individuals at risk, with most studies focussing on the impact of individual demographics on mRNA vaccine efficacy. There is also a lack of studies assessing individuals’ determinants on cellular versus humoral immunity.

Here we assessed a population-based cohort from the COVIDENCE study^16^, who had either two doses of BNT162b2 or ChAdOx-n-CoV-19 to determine the influence of individual demographic variables such as age, sex, BMI, and pre-vaccination IgG/A/M anti-Spike seropositivity on SARS-CoV-2 antibody and antigen-specific CD4+ and CD8+ T cell response, bulk T cell functional phenotypes and antigen-induced whole blood cytokine secretion following to the first two doses of the vaccines BNT162b2 and ChAdOx-n-CoV-19.

## 2. Materials and Methods

### 2.1 Study design

This study was a sub-study nested within the CORONAVIT randomised controlled trial as reported elsewhere^16, 17^. 6200 U.K. residents aged 16 years or older participated in the COVIDENCE U.K. study as described previously^16^, all of whom provided dried blood spot samples (prior to vaccination) for the determination of combined IgG, IgA and IgM (IgG/A/M) antibody responses to the Spike protein of SARS-CoV-2, as described below. A subset of the participants (123 individuals in total) returned after COVID-19 vaccination and provided a dried blood spot for antibody analysis and a sodium heparin blood sample to assess cellular immunity to SARS-CoV-2 using antigen-stimulated whole blood and peripheral blood mononuclear cell (PBMC) assays. Serum was also collected for C Reactive protein (CRP) and SARS-CoV-2 neutralising antibody quantification. The trial was sponsored by Queen Mary University of London, approved by the Queens Square Research Ethics Committee, London, U.K. (ref 20/HRA/5095) and registered with ClinicalTrials.gov (NCT04579640) on 8 October 2020, before enrolment of the first participant on 28 October 2020.

### 2.2 *In vitro* Cellular Immune Response Assays

Heparinised blood collected from all sub-study participants was used to assess cellular immunity post vaccination using two approaches:

Whole blood was stimulated in the presence or absence of PepTivator® SARS-CoV-2 Prot_S Complete (1µg/mL; Miltenyi Biotec) or E. coli lipopolysaccharide (LPS, 1–1000 ng/mL; Invivogen) for 24 h at 37°C in 5% CO_2_. Following incubation plasma was collected and stored at -80°C for cytokine assessment by cytometric bead array as detailed below.

PBMCs were isolated from heparinised blood using Ficoll (Merck Life Science) density gradient, washed twice in Hanks Balanced Salt Solution (Merck Life Science) and cryopreserved in 10% dimethylsulfoxide (DMSO) in Fetal Calf Serum (Invitrogen).

Subsequently, cryopreserved PBMCs were recovered and stimulated with nothing (negative control) or PepTivator® SARS-CoV-2 Prot_S Complete (Miltenyi Biotec, 1µg/mL) or 1µg/mL of soluble CD3 monoclonal antibody (OKT3, Functional Grade, Invitrogen) for 1 h at 37°C in 5% CO_2_. Brefeldin A (2.5 µg/mL) was then added to the cells, which were incubated for a further 15 h at 37°C in 5% CO_2_. Cells were collected and analysed by flow cytometry as detailed below.

### 2.3 Flow Cytometric Analysis

Cells were cell surface stained for CD3 (HIT3a), CD4 (RPA-T4), CD8 (SK1), CD27 (O232), CD45RA (HI100) and Zombie NIRTM viability dye (Biolegend, San Diego, CA, USA) in the presence of Brilliant Buffer (BD Biosciences). Cells were washed and then fixed in Intracellular Fixation Buffer (eBioscience), permeabilised in eBioscience Permeablization Buffer and stained for intracellular IL-2 (JES6-5H4), IFN-γ (4S.B3) and TNF (Mab11, Biolegend). Cells were then washed and acquired using the ACEA Novocyte 3000 flow cytometer (Agilent). Data were analysed using FlowJo Version X (BD Biosciences). A representative flow cytometry gating strategy is shown in Supplementary Figure 1. Following single cell and live cell gating, once CD3+ CD4+ and CD8+ cells were identified then proportion cytokine expression was calculated in the stimulated conditions based upon the unstimulated control. In addition, T cell phenotypic staining was performed on unstimulated cells using the markers CD27 and CD45RA to determine if the CD4+ or CD8+ T cell are naïve (CD27+CD45RA+), central memory (CM, CD27+CD45RA-), effector memory (EM, CD27-CD45RA-) or senescent-like effector memory re-expressing CD45RA (EMRA; CD27- CD45RA+). A representative flow cytometry gating strategy for T cell memory phenotypes and representative plots from young (<40 years) and old (≥ 60 years) donors is shown in Supplementary Figure 2.

### 2.4 Antibody analysis

#### 2.4.1 Anti-Spike (S) IgG/A/M serology testing

Anti-S IgG/A/M titres were determined by the Clinical Immunology Service at the Institute of Immunology and Immunotherapy of the University of Birmingham using an ELISA that measures combined IgG/A/M responses to the SARS-CoV-2 trimeric Spike glycoprotein (product code MK654, The Binding Site [TBS], Birmingham, U.K.), as previously described^15^. This assay has been CE-marked with 98.3% (95% confidence interval [CI] 96.4–99.4) specificity and 98.6% (92.6–100.0) sensitivity for RT-PCR-confirmed mild-to-moderate COVID-19 ^18^, and has been validated as a correlate of protection against breakthrough SARS-CoV-2 infection in two populations^18, 19^. A cut-off ratio relative to the TBS assay cut-off calibrators was determined by plotting 624 pre-2019 negatives in a frequency histogram. A cut-off coefficient was then established for IgG/A/M (1.31), with ratio values classed as positive (≥1) or negative (<1). Dried blood spot eluates were pre-diluted 1:40 with 0.05% PBS-Tween using a Dynex Revelation automated absorbance microplate reader (Dynex Technologies). Plates were developed after 10 min using 3,30,5,50-tetramethylbenzidine core, and orthophosphoric acid used as a stop solution (both TBS). Optical densities at 450 nm were measured using the Dynex Revelation.

#### 2.4.2 SARS-CoV-2 Neutralising Antibody

Serum titres of neutralising antibodies to SARS-CoV-2 were measured as previously described using an authentic virus (Wuhan Hu-1 strain) microneutralisation assay^20^.

### 2.5 C-Reactive Protein Assessment

Serum was analysed for C-Reactive protein concentrations using Human C-Reactive Protein/CRP DuoSet ELISA (Biotechne, R+D systems) according to the manufacturers protocol.

### 2.6 Cytometric Bead Array

A cytometric bead array to measure IL-8, IL-6, IFN-γ and TNF in whole blood stimulated plasma was carried out according to the manufacturer’s protocol (BD Biosciences). Samples were analysed using the ACEA Novocyte 3000 flow cytometer (Agilent). The lower limit of detection was 1.5 pg/mL.

### 2.7 Analysis

Principle component analysis (PCA) and general linear modelling (GLM) was conducted using Qlucore Omics Explorer 3.7 (Qlucore AB, Lund, Sweden). Analyte concentrations were log2 converted and normalised to the mean for each analyte with variance -1 to +1. Missing values were imputed by K nearest neighbours (k-NN). Of the 127 participants with acquired data, 12 were excluded from statistical analysis (two due to haematological malignancy with an absence of CD4+ T cells, four due to immunodeficiency, three for being on immunosuppressive medication, one as having reported worst health and two who were identified as outliers on PCA (Supplementary Figure 3). Parameters whose concentration differed significantly between vaccination type, sex and pre-vaccination anti-Spike serostatus were identified using the t-test for GLM initially without and then with adjustment for covariates identified in univariate analysis to influence the inflammatory profile, with an additional adjustment included for COVIDENCE trial arm allocation [placebo, low vitamin D supplementation, high vitamin D supplementation] although we found no significant effect of vitamin D supplementation on vaccine efficacy in the trial^16^. Covariate adjustment was performed using the eliminated factors approach that fits a multiple regression model to all covariates and subtracts the expression values predicted by this model from the observed values in order to remove covariate effects between patients^21^. Differences based on BMI category, ethnicity and general health category were assessed by F- test for GLM with similar adjustment approach for covariates. Parameters affected by age, BMI value and days post second vaccination were identified using a liner regression model, whilst anti-spike ratios pre- and post-vaccination, NAB titres post-vaccination and inter-vaccine days were analysed by quadratic regression, with similar covariate adjustment approach. These analyses yield t-statistics representing the magnitude of difference in concentration of a given parameter between groups being compared (calculated as the regression co-efficient for each parameter divided by its standard deviation), p values and q values (Benjamini-Hochberg false discovery rate (FDR). Thresholds of 0.05 were applied for p and q values throughout.

## 3. Results

### 3.1 Identification of individual correlates of immunogenicity

Characteristics of the 115 participants with humoral and cellular data included in the analyses are presented in Table 1. The average age was 66.4 years (IQR 61.0-68.9), 47 (41%) males and 68 (59%) females, 57 (49.6%) had a BMI less than 25, 45 (39.1%) had a BMI of 25-30 and a further 13 (11.3%) had a BMI of greater than 30. Of the participants 77 (66.9%) received ChAdOx1 and 38 (33.0%) received BNT162b2. To identify demographic and technical factors associated with humoral and cellular responses to COVID-19 vaccination we first performed univariate analysis of ten factors: age, sex, ethnicity, general health category, vaccine type, number of days between 1st and 2nd vaccination [inter- vaccine days], days post second vaccine, pre-vaccine SARS-CoV-2 sero-status, BMI value, and BMI category. This analysis identified seven factors which had a significant association with cellular and/or humoral immune responses: age, sex, vaccine type, inter-vaccine days, days post second vaccine, pre-vaccine SARS-CoV-2 sero-status and BMI category (Supplementary Tables 1a-i). These seven factors were included in subsequent multivariate analyses to identify independent associations.

**Table 1:** Participant Characteristics (n=115) Abbreviations: IQR, inter-quartile range; s.d., standard deviation; Ig, Immunoglobulin. Self-assessed general health is in 115 participants, and median time from date of second vaccine was measured in 112 participants.

| Characteristic |  |  |
| --- | --- | --- |
| Age | Median age, years (IQR) | 66.4 (61.0-68.9) |
|  | Age range, years | 23.5–78.6 |
| Sex, N (%) | Male | 47 (40.9%) |
|  | Female | 68 (59.1%) |
| Ethnicity, N (%) | White | 107 (93.0%) |
|  | South Asian | 1 (0.9%) |
|  | Black/African/Caribbean/<br>Black British | 0 |
|  | Mixed/Multiple/Other | 7 (6.1%) |
| Body mass index, kg/m<sup>2</sup>, N (%) | <25 | 57 (49.6%) |
|  | 25-30 | 45 (39.1%) |
|  | >30 | 13 (11.3%) |
| Highest educational level attained, N (%) <sup>3</sup> | Primary/Secondary | 9 (7.8%) |
|  | Higher/Further (A levels) | 9 (7.8%) |
|  | College | 55 (47.8%) |
|  | Post-graduate | 42 (36.5%) |
| Quantiles of index of multiple deprivation, N (%) | Q1 (most deprived) | 19 (16.5%) |
|  | Q2 | 21 (18.3%) |
|  | Q3 | 38 (33.0%) |
|  | Q4 (least deprived) | 37 (32.2%) |
| Tobacco smoking, N (%) | Non-current/never smoker | 111 (96.5%) |
|  | Current smoker | 4 (3.5%) |
| Alcohol consumption/week, units, N (%) | None | 21 (18.3%) |
|  | 1-7 | 43 (37.4%) |
|  | 8-14 | 26 (22.6%) |
|  | 15-21 | 16 (13.9%) |
|  | 22-28 | 8 (7.0%) |
|  | >28 | 1 (0.9%) |
| Self-assessed general health, N (%) | Excellent | 29 (25.4%) |
|  | Very good | 43 (37.7%) |
|  | Good | 31 (27.2%) |
|  | Fair | 11 (9.6%) |
|  | Poor | 0 |
| Pre-vaccination anti-S IgG/A/M serostatus, N (%) | Negative | 100 (81.7%) |
|  | Positive | 15 (12.2%) |
| Type of vaccine received for primary course, N (%) | 2 x ChAdOx1 | 77 (66.9%) |
|  | 2 x BNT162b2 | 38 (33.0%) |
| Median inter-dose interval, weeks (IQR) |  | 11.0 (10.0-11.3) |
| Median time from date of second vaccine dose to date of sampling, weeks (IQR) |  | 11.9 (10.0-14.6) |

### 3.2 Immune correlates with anti-Spike and SARS-CoV-2 neutralising antibodies after SARS-CoV-2 vaccination

First, we investigated whether there was any between humoral and cellular immune responses we measured. Post-vaccination titres of anti-Spike combined IgG/A/M antibody ratio responses were identified in multivariate analysis to be associated with neutralising antibody concentrations, three spike-specific CD4+ T cell phenotypes and spike-stimulation induced IFNγ secretion (Figure 1, Supplementary Table 2). In addition, neutralising antibody concentrations significantly correlated with SARS-CoV-2-specific TNF+ CD8+ T cell frequency (p = 0.0003, Supplementary Table 3) As expected, post-vaccination titres of neutralising and anti-Spike IgG/A/M antibody ratio correlated positively with each other (R = 0.47, p<0.0001, Figure 1A). The three spike-specific CD4+ T cell populations from PBMC SARS-CoV-2 peptide stimulated cultures that positively correlated with anti-Spike IgG/A/M antibody ratio post-vaccination were all double positive cytokine producers: IFN-γ+IL2+ (R = 0.36, p = 0.004); TNF+IL-2+ (R = 0.35, p =0.0004); IFN-γ+TNF+ (R=0.32, p = 0.001) (Figure 1C). In addition, anti-S IgG/A/M antibody ratio correlated positively with IFNy secretion from SARS-CoV-2 peptide-stimulated whole blood (R = 0.31, p = 0.001; Figure 1B). There was also a trend (q = 0.1) for a positive correlation between anti-Spike IgG/A/M and SARS-CoV-2 peptide-specific CD4+ IL-2 single positive T cells (Supplementary Table 2).

**Figure 1:**
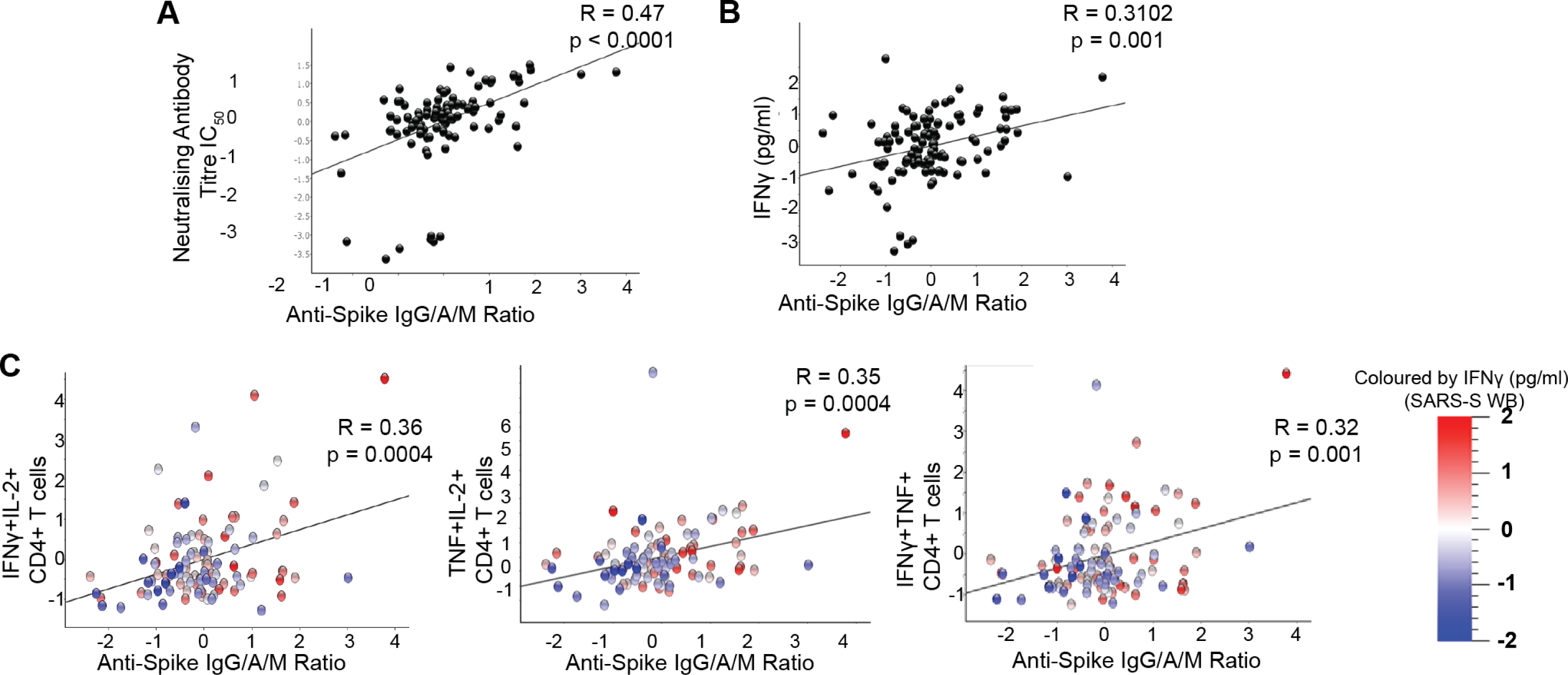
Humoral and cellular correlates of anti-S IgG/A/M antibody ratio after COVID-19 vaccination. Correlation between post-COVID-19 vaccine anti-S IgG/A/M antibody ratio and **A,** neutralising antibody titre IC_50_, **B,** whole blood IFNγ production after S peptide stimulation and **C,** percent of cytokine positive CD4+ T after PBMC stimulation with S peptide as determined by intracellular cytokine staining. Coloured according to IFNy production from S peptide stimulated whole blood (SARS-S WB). Data presented is normalised including log2 transformation and adjusted for the covariates.

### 3.3 Influence of pre-vaccination anti-S IgG/A/M antibody ratio on post-vaccine humoral and cellular responses

We next determined whether there was a relationship between pre-vaccine anti-Spike IgG/A/M antibody ratio on post-vaccine humoral and cellular responses (Supplementary Table 4). Those who were considered seropositive pre-vaccination (anti-Spike IgG/A/M antibody ratio ≥1) had a significantly higher frequency of IFNγ+ CD4+ and CD8+ SARS- CoV-2-specific T cells (Figure 2A and B) post-vaccination. There was also a significant correlation with pre-vaccine anti-S IgG/A/M antibody ratio and post-vaccine frequency of effector memory CD4+ T cells in unstimulated PBMC (p = 0.006; Supplementary Table 4). Together, these data demonstrate that those seropositive due to prior SARS-CoV-2 infection had a stronger cellular immune response to SARS-CoV-2 vaccination than those who did not.

**Figure 2:**
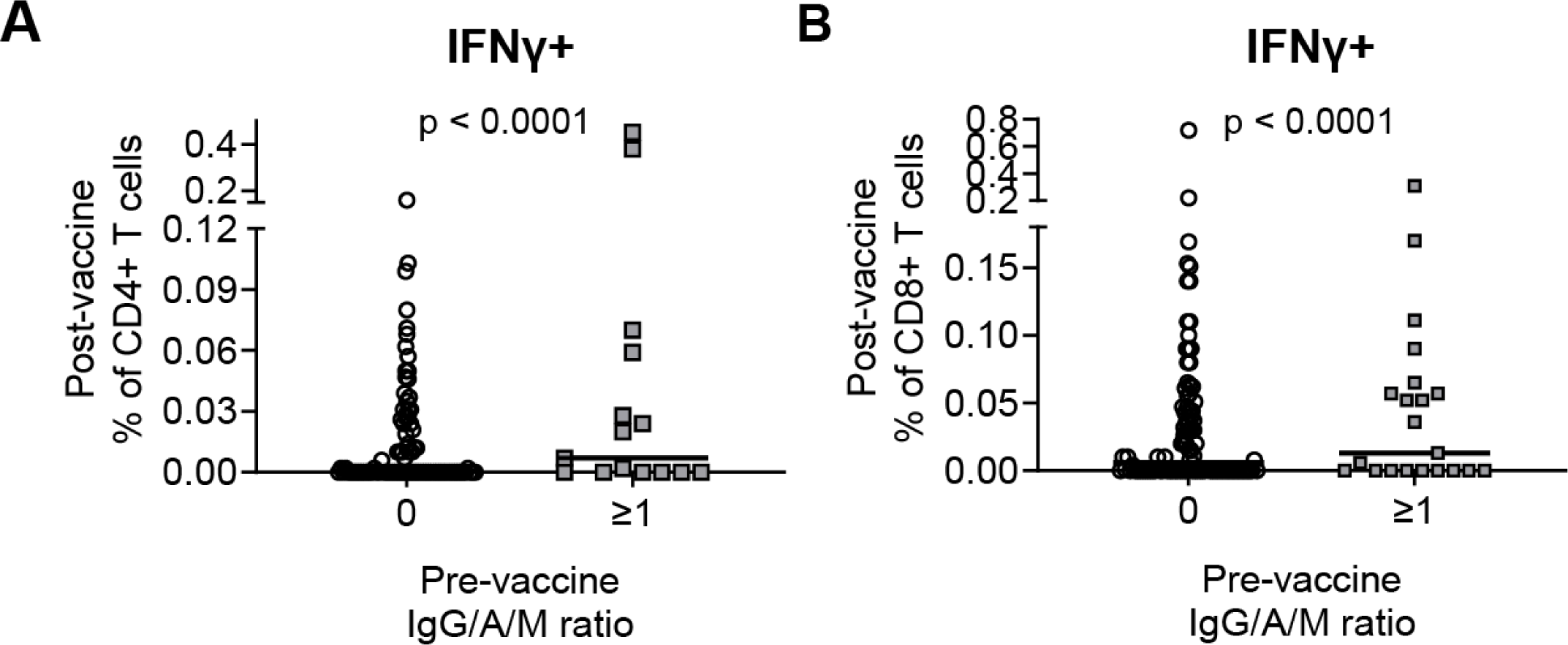
Humoral and cellular responses post-vaccination stratified by pre-vaccination SARS-CoV-2 serostatus. Cumulative data showing frequency of IFNy+ **A,** CD4+ and **B,** CD8+ T cells after S peptide stimulation stratified by pre-vaccine anti-S IgG/A/M antibody ratio as a measure of negative (0) or positive (≥1) serostatus. Median indicated by line.

### 3.3 Influence of vaccine type, inter-dose interval and time from vaccination to sampling on post-vaccine humoral immune responses

We found that there were significantly higher anti-S IgG/A/M antibody ratio and neutralising antibody titres in those that received BNT162b2 as compared to ChAdOx1-nCoV-19 (Figure 3A and B). Interestingly, however, there was no significant difference in unstimulated or antigen-stimulated cellular responses that we measured in PBMC or whole blood between those who received BNT162b2 compared with those who received ChAdOx1-nCoV-19 (Supplementary Table 5). When analysing whether there was an impact of the number of days between vaccinations and measured immune responses, we only identified a trend for the frequency of SARS-CoV-2 peptide-specific TNF+IFNγ+ CD4+ T cells and TNF+ CD8+ T cells (p ≤ 0.007; q=0.16) on multivariate analysis (Supplementary Table 6). However, we did find that the delay from the date of the second vaccine dose to the date of blood draw was positively correlated with the level of SARS-CoV-2 peptide-induced whole blood secretion of IL-6, IL-8 and TNF (p ≤ 0.0004), and a trend for negative correlation with Neutralising antibody titres (p = 0.01; q = 0.14) (Supplementary Table 7).

**Figure 3:**
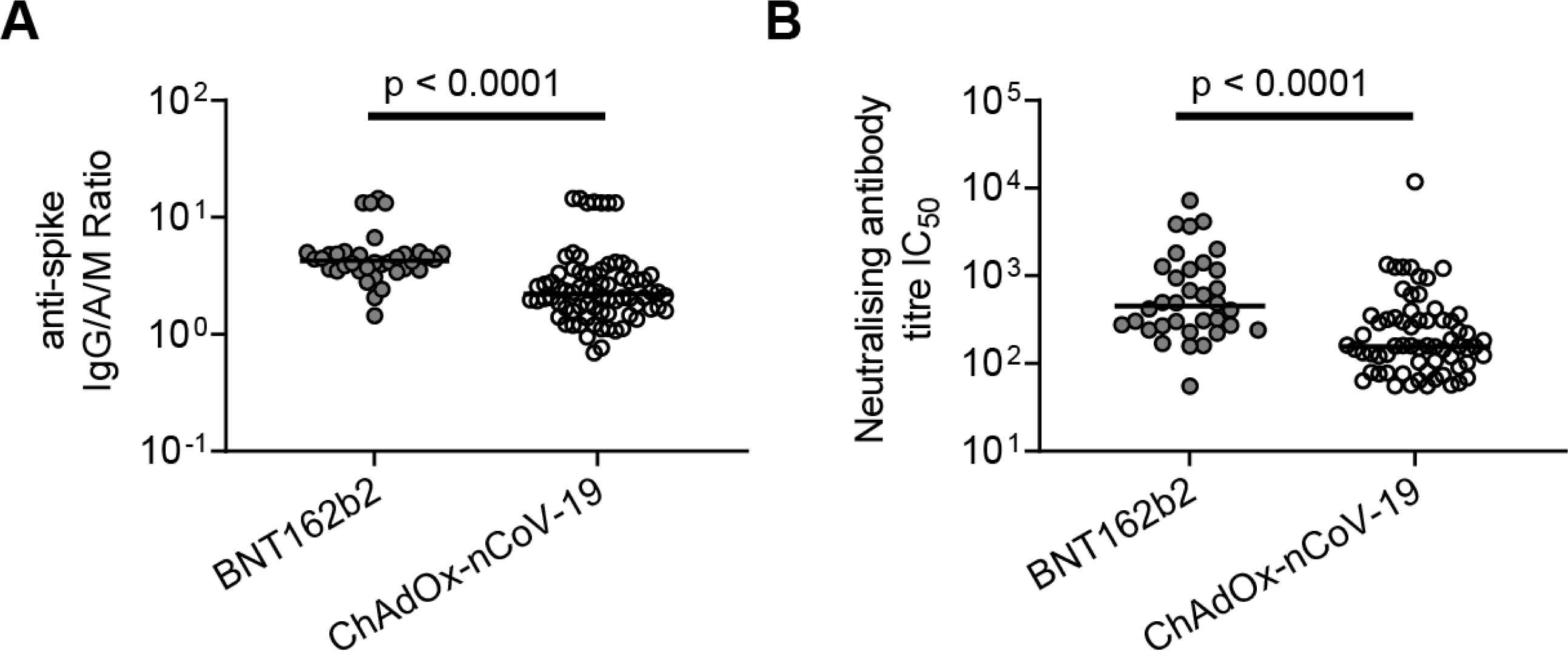
Post-vaccination antibody titres by vaccine type. Cumulative data showing **A,** anti-Spike IgG/A/M antibody ratio and **B,** neutralising antibody titre IC_50_ in participants post-vaccination with either BNT162b2 or ChAdOx1-nCoV-19. Line indicated Median.

### 3.4 Increasing age significantly altered the immune response to vaccination

Having determined the vaccine-type and timing variables associated independently associated with vaccine induced humoral and cellular immune responses, we next analysed demographic correlates adjusting for these in the multivariate analysis. We found that increasing age was independently associated with lower anti-S antibody titres post-vaccination (Figure 4). The frequency of cytokine positive CD4+ T cells (double positive for IFN-γ+IL-2+ [p=0.012] or TNF+IL-2+ [p=0.012]) from PBMC SARS-CoV-2 peptide stimulated cultures post-vaccination also negatively correlated with increasing age (Supplementary Table 8). As would be expected, we also found increasing age was independently associated with lower frequency of naïve CD8+ T cells (p < 0.0001) and higher frequency of CD8+ EM (p = 0.004) and EMRA (p=0.036). These data collectively show that increasing age was associated with reduced humoral immunity associated with reduced double-positive cytokine producing spike-specific CD4+ T cells.

**Figure 4:**
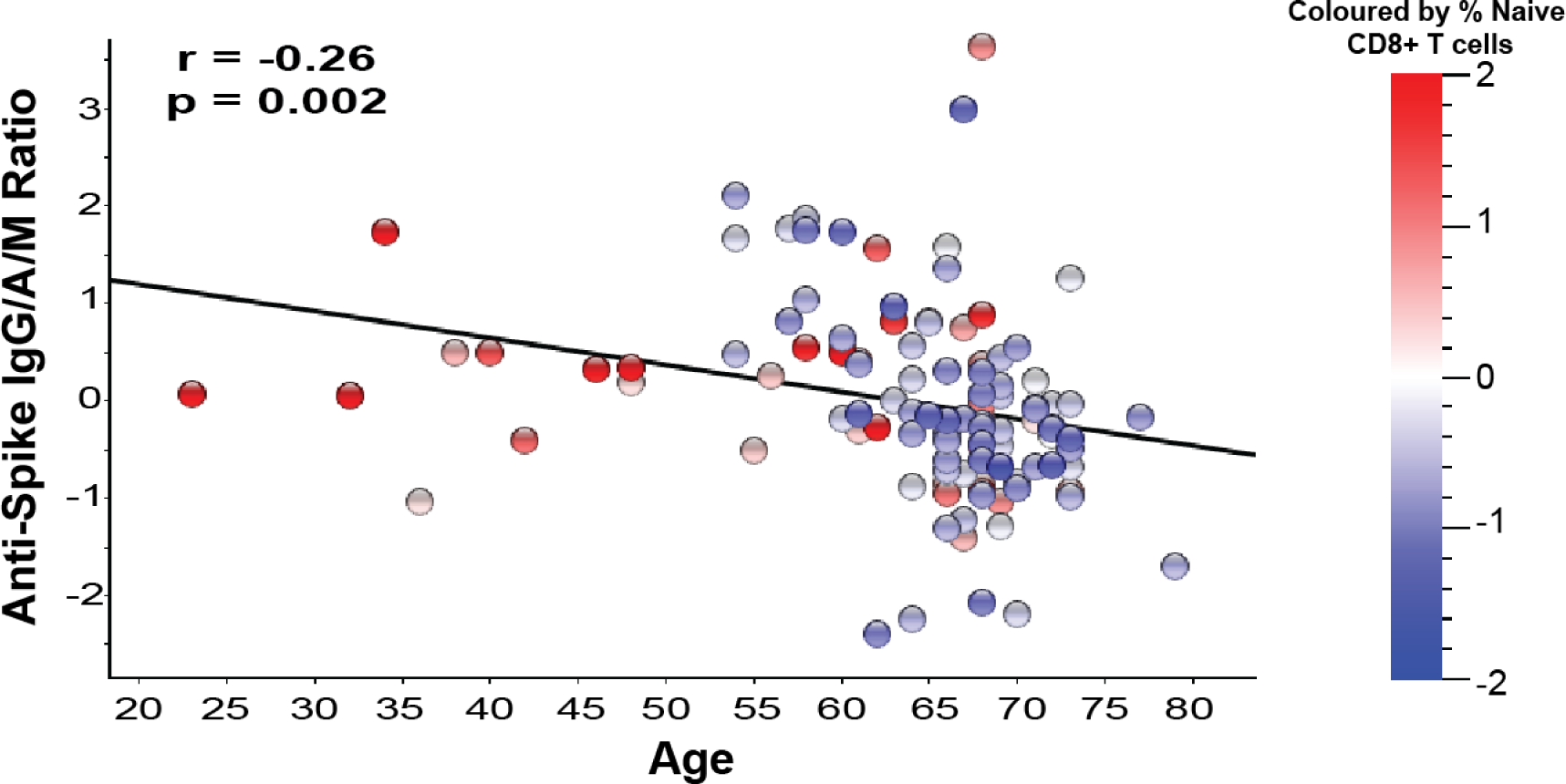
Relationship between post-vaccination anti-S IgG/A/M antibody ratio, percent naïve CD8+ T cells and age Correlation between age and post-vaccine anti-Spike IgG/A/M antibody ratio. Coloured according to frequency of naïve (CD45RA+CD27+) CD8+ T cells present in the peripheral blood of the individuals. Data presented is normalised including log2 transformation and adjusted for the covariates.

### 3.5 Influence of BMI and gender on post-vaccine immune responses

Finally, we tested for associations between sex and BMI and SARS-CoV-2 cellular and humoral immunity. We found a highly significant lower frequency of SARS-CoV-2-specific TNF+ CD4+ and CD8+ T cells (p < 0.0001) correlated with increasing BMI category (Supplementary Table 9), as well as trends of lower SARS-CoV-2-specific IL-2+ CD4+ T cells and higher CRP (p ≤ 0.014, q < 0.156). There was no significant association between higher BMI category and anti-Spike or neutralising antibodies, despite our finding that the level of neutralising antibodies significantly correlated with SARS-CoV-2-specific TNF+ CD8+ T cells which were decreased with increasing BMI.

When analysing associations between sex immune correlates, the most significant difference was in our control assay with higher LPS induced IL-6 secretion in whole blood stimulated plasma (p < 0.0001). Males also had a higher frequency of CD8+ (p = 0.0001) and CD4+ (p = 0.011) EM T cells, whilst females had a higher frequency of naïve CD4+ (p = 0.012) and CD8+ (p = 0.002) T cells (Supplementary Table 10). However, these differences did not impact upon SARS-CoV-2 specific cellular or humoral responses post-vaccination between males and females with no significant differences observed (Supplementary Table 10). Collectively these data show sex had no impact on SARS-CoV-2 vaccine immunogenicity, whilst BMI had significant effects on single cytokine producing SARS-COV- 2-specific T cell functions we independently identified to be associated with SARS-CoV-2 neutralising antibody concentrations post-vaccination.

## 4. Discussion

In this study, we observed that there was a significant positive correlation between humoral and cellular SARS-CoV-2-specific immunity post vaccination which were modified by increasing age and BMI but not sex of individuals. The levels of anti-Spike IgG/M/A and SARS-CoV-2 neutralising antibodies were significantly correlated with SARS-CoV-2 induced cytokine secreting double positive CD4+ T cells or TNF+ CD8+ T cells, respectively. As expected, those individuals who were seropositive prior to vaccination had the largest humoral and cellular immune response following SARS-CoV-2-vaccination, irrespective of vaccine type. We observed that there were significantly higher antibody titres in participants that received BNT162b2 as compared to those that received ChAdOx1-nCoV-19. However, no difference was observed in antigen-specific cellular immunity post-SARS-CoV-2 vaccination that explained these vaccine immunogenicity differences. By contrast, no impact of biological sex was observed for SARS-CoV-2 vaccine cellular and humoral immunity, despite cellular functional phenotype differences identified.

Previous studies have observed that BNT162b2 was more efficacious as a vaccine as compared to ChAdOx1-nCoV-19 at reducing infections and hospital admissions^22, 23, 24^. We observed that there were increased anti-S IgG/A/M antibody ratio and neutralising antibody titres with BNT162b2 as compared to ChAdOx1-nCoV-19, in line with previous observations^11, 15^. Interestingly, we did not observe any association between vaccine type on SARs-CoV-2 antigen-specific T cell immunity. However, it is important to note that both vaccines still resulted in a good humoral and cellular immune response in the participants in this study. In line with the current literature^15, 25^, we observed that there was increased COVID-19 vaccine immunogenicity in individuals that have had a prior infection with SARS-CoV-2. This boost in immunogenicity is believed to be due to the phenomenon of hydrid vigor immunity^26^.

It has been shown that older adults have reduced vaccine efficacy to influenza and shingles vaccine^27, 28^, which is believed to be in part to immunosenescence and inflammageing^29, 30^. This was particularly worrying at the start of the pandemic as older adults had increased morbidity and mortality from COVID-19^8^. Therefore, when the vaccines were developed, studies were performed to assess COVID-19 vaccine efficacy in older adults – the resulting studies were conflicting with results demonstrating similar vaccine efficacy (as determine by infection events and hospitalization) by age or reduced vaccine efficacy. In this study we observed that increasing age was associated with reduced post-vaccine anti-S antibody titre, in line with other studies which have shown reduced responses post-SARS-CoV-2 vaccination^10, 31^, although older age has been associated with lower risk of breakthrough infection^24^. This reduced antibody response with increasing age has been shown to be alleviated after SARS-CoV-2-vaccine booster ^9, 31^. Interestingly, we also observed a trend for decreased IFN-γ+IL-2+ and TNF+IL-2+ CD4+ T cells which we found to be independently associated with post-vaccination anti-Spike IgG/M/A levels. This therefore links decreased T cell immunity with age to decreased humoral immunity.

Due to the inflammatory nature of obesity^32^, and the detrimental role that inflammation plays in antigen-specific immunity^29^, it has been proposed that obesity could result in worse vaccine efficacy and immunogenicity. Unlike in the large COVIDENCE cohort where we found increased BMI associated with higher anti-Spike IgG/A/M antibody ratio titres^15^ we did not observe the same relationship in this sub study. Interesting, despite not identifying any relationship to increased BMI with humoral immunity, BMI category was associated with SARS-CoV-2 specific TNF+ CD8+ and CD4+ T cells, with TNF+ CD8+ T cells independently highly correlated with SARS-CoV-2 neutralising antibody titres. It may be that a non-linear relationship exists between BMI category and neutralising antibodies. We also may not have power to detect this relationship, given we also did not identify the relationship between BMI and anti-Spike IgG/A/M antibody ratios identified in the larger cohort which included more individuals with BMI > 30 (11.8% here vs 18.5% in larger cohort). However, other studies have also either not observed any effect of increased BMI on SARS-CoV-2-vaccination using BNT162b2 ^10, 12^ or showed reduced antibody response post- BNT162b2 or CoronaVac vaccination in individuals with high BMI^33^. The difference between these studies may be due to the lower average BMI observed in our study.

The main strength of this study was that it was a population-based study in which we applied multivariate analyses to identify independent and associated predictors of immune correlates post-COVID-19 vaccination. In addition, most studies to-date have focussed on BNT162b2 vaccine immunogenicity, the strength of our study is that we assessed vaccine Pfizer BNT162b2 and ChAdOx1-nCoV-19 vaccination allowing us to compare immune responses to the two vaccines administered at the same stage of the pandemic. One finding from our study was that we observed that the concentration of anti-Spike IgG/A/M antibody ratio significantly correlated with antigen-specific CD4+ T cell immunity and induced cytokine secretion – demonstrating the strengths of combining a comprehensive characterization of humoral responses with PBMC and whole blood antigen stimulation assays. The limitations to this study were that we had a bias towards older individuals in the study population, with limited number of people under 35 years old. In addition, there was a modest bias towards female sex (59% vs 41%) in our study population. Another limitation to the study was that it was a single time-point after COVID-19 vaccination, this means we were not able to look at waning immunity over time.

To conclude age, vaccine type and prior infection influence the humoral and cellular immune response to SARS-CoV-2 vaccination, whilst BMI affected cellular immunogenicity that independently associated with neutralising antibody levels. We found no difference in vaccine immunogenicity by sex. This study supports the need to include diverse populations during early vaccine efficacy testing, with demographic features related to poor infection outcome, and the benefits of parallel measurement of humoral and cellular immune functions to enable a thorough understanding of vaccine induce immune correlates of protection.

## Data Availability

All data produced in the present work are contained in the manuscript

## Acknowledgements

We would like to thank Professor Áine McKnight whose lab developed the neutralizing antibody assay that was used in this study. We thank all participants in COVIDENCE UK and the following organizations that supported study recruitment: Asthma UK and the British Lung Foundation, the British Heart Foundation, the British Obesity Society, Cancer Research UK, Diabetes UK, Future Publishing, Kidney Care UK, Kidney Wales, Mumsnet, the National Kidney Federation, the National Rheumatoid Arthritis Society, the North West London Health Research Register (DISCOVER), Primary Immunodeficiency UK, the Race Equality Foundation, SWM Health, the Terence Higgins Trust, and Vasculitis UK.

## Author contributions

ARM wrote the study protocol. ESC designed the cellular assays and along with WC performed the whole blood and PBMC experiments S peptide– and LPS experiments. SF and AGR developed, validated, and performed assays for anti-spike immunoglobulin (Ig) G, IgA, and IgM (IgG/A/M) Antibody ratio. NP and WL were responsible for patient recruitment and sample collection. JMG, CP, and AM developed, validated, and performed assays for neutralizing antibodies. AKC and GV contributed to data management and statistical analyses. ESC and AKC generated figures. ESC, AKC and ARM wrote the first draft of the report. All authors reviewed the manuscript critically for important intellectual content, gave final approval of the version to be published.

## Supplementary Figures

**Supplementary Figure 1:**
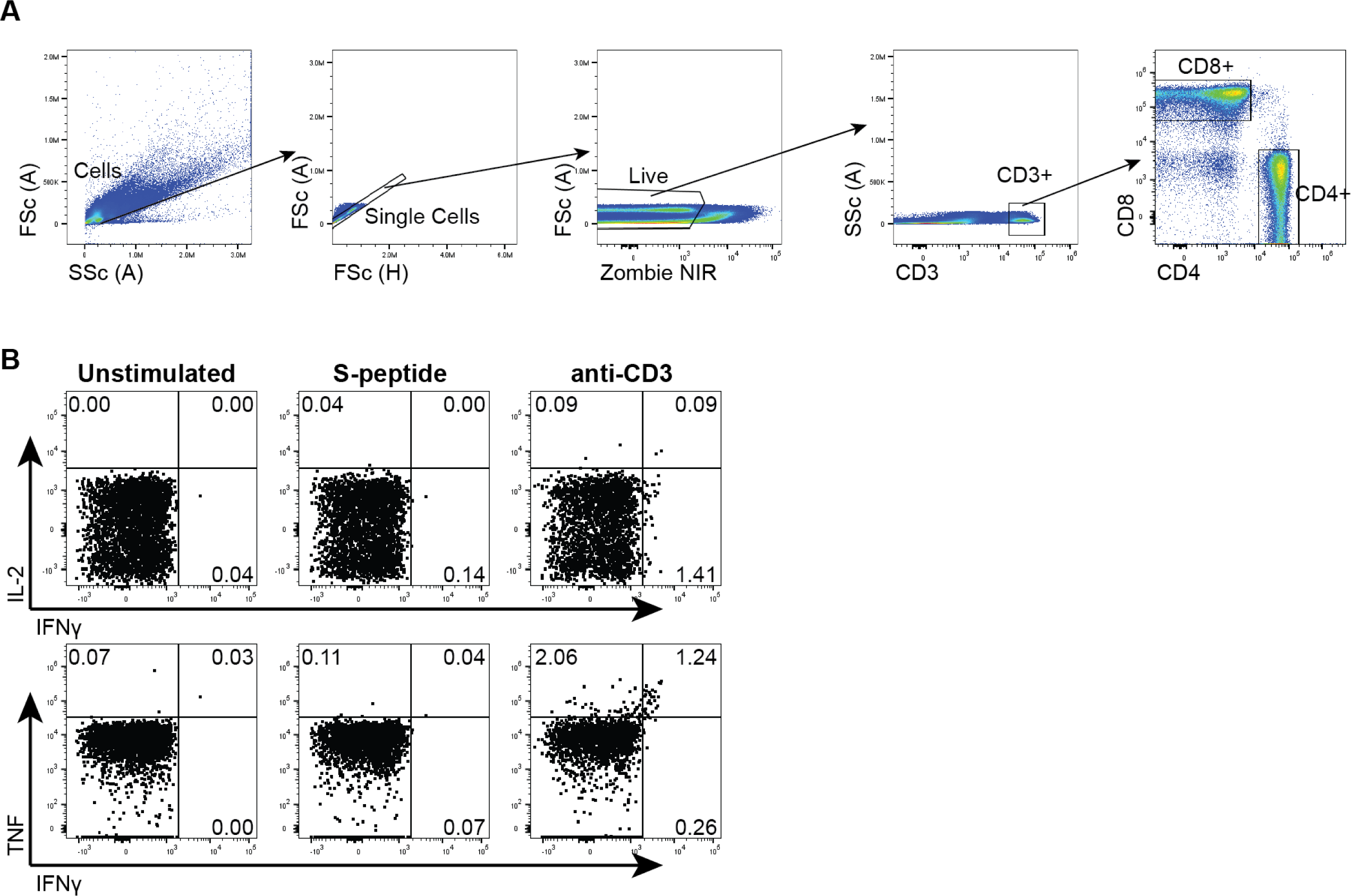
Representative flow cytometry gating strategy to identify T cells and intracellular cytokines. PBMCs were assessed by cell surface and intracellular flow cytometry analysis. **A,** Representative gating strategy to determine CD4+ and CD8+ T cells. Lymphocytes were identified from Forward (FSc) and Side (SSc) scatter plots, subsequently single cells were identified. Then Live cells were selected as being Zombie Near Infrared (NIR) Live dead stain negative, then T cells were identified based upon CD3 expression. Finally CD4+ and CD8+ T cells were identified within the CD3 gate. **B,** Representative IFNy, IL-2 and TNF staining is shown of unstimulated (negative control), S-peptide stimulated and soluble anti-CD3 stimulated (positive control) conditions.

**Supplementary Figure 2:**
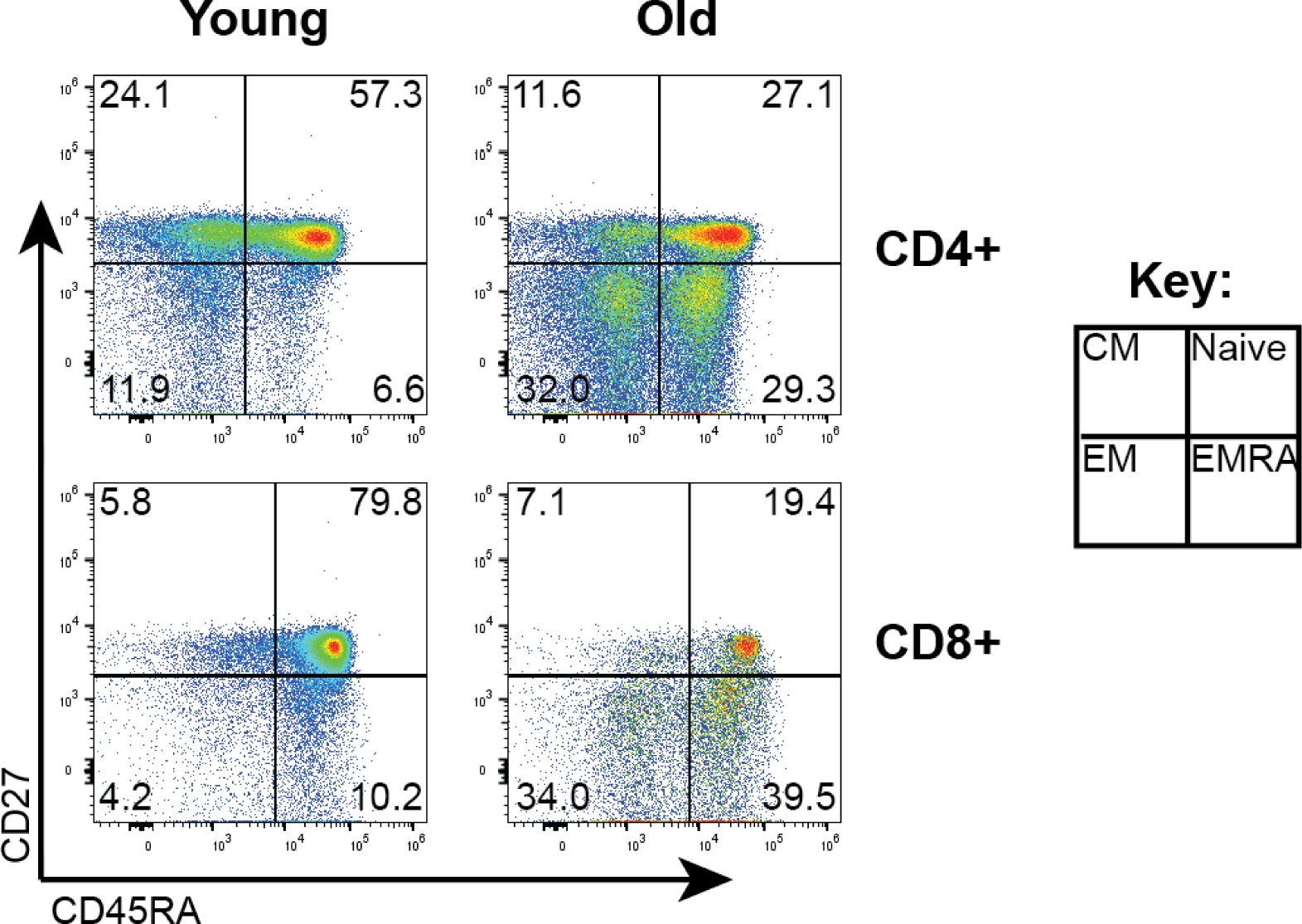
Representative gating strategy for phenotyping of T cells. CD4+ and CD8+ T cells were identified within the CD3 gate, as shown in supplementary Figure 1. Representative CD45RA and CD27 staining shown in CD4+ and CD8+ T cells from a younger (<40 years) and older (≥65 years) person from unstimulated PBMCs. CD45RA+CD27- are considered naïve, CD45RA-CD27+ are considered Central memory (CM), CD45RA-CD27- Effector memory (EM) and CD27-CD45RA+ are senescent-like T cells known as Effector Memory re-expressing CD45RA (EMRA).

**Supplementary Figure 3:**
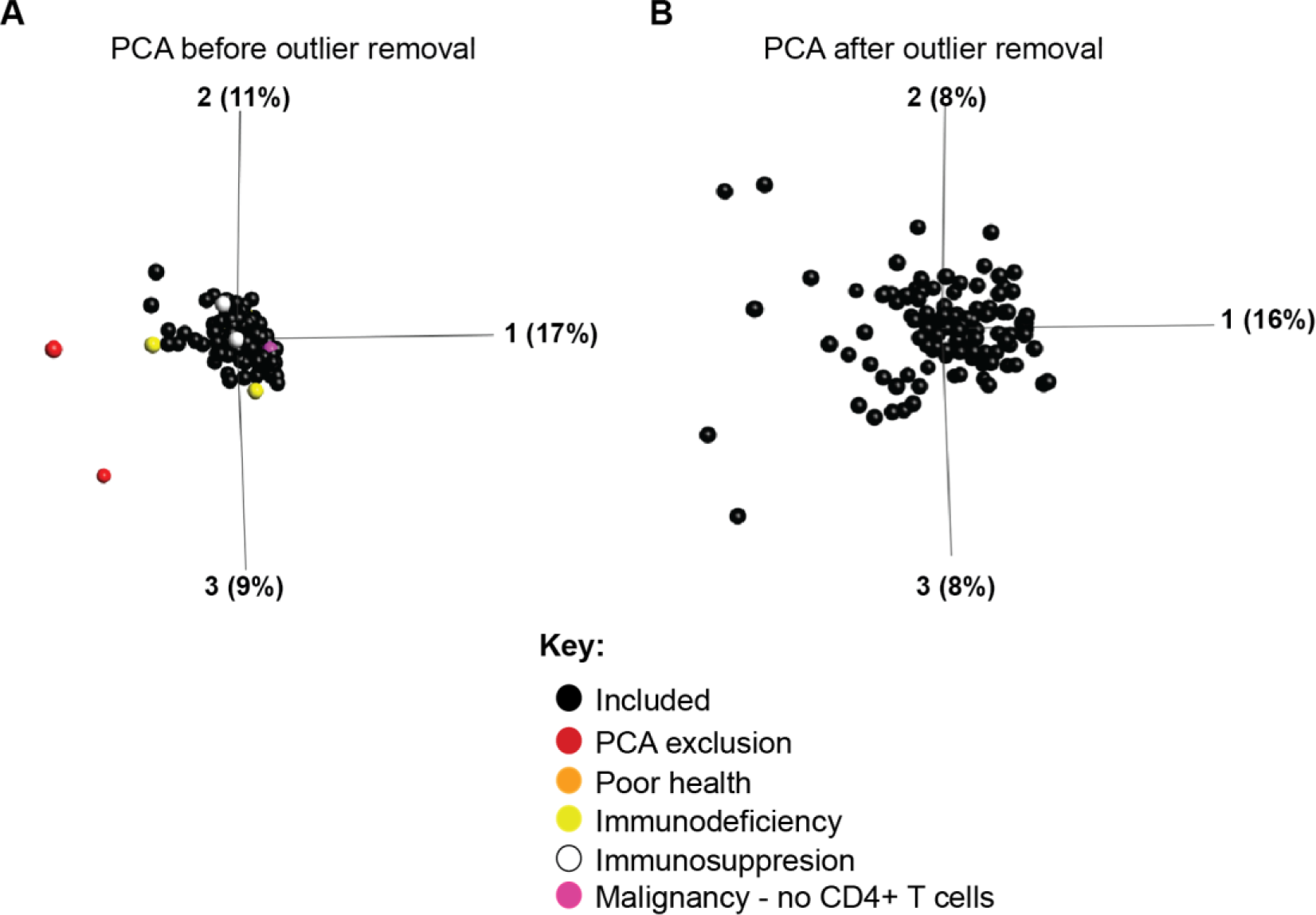
Principal component analysis (PCA) before and after outlier removal. **A,** Twelve samples were excluded in total. Two samples were identified based upon principle component analysis (PCA) as being outliers and were removed (red). Additionally samples were excluded due to immunosuppressive medication (white; n=3) or immunodeficiency (yellow; n=4) or poorest general health (orange, n=1) or as having malignancy and absence of CD4+ T cells (as determined by flow cytometry; pink; n=2). **B,** PCA plot of all included samples (n=115; black) used for analyses showing good separation of samples across the first three components.

