## Supplementary Tables 2-10 for "Influence of age, sex, body habitus, vaccine type and anti-S serostatus on cellular and humoral responses to SARS-CoV-2 vaccination"

(see separate excel sheet for supplementary Tables 1a-i)

**Supplementary Tables 1a-i: Factors with analytes sigificant at q-value <0.15 on univariate analysis were included in multivariate anlaysis.**

BMI category was selected over BMI value. Neut Ab = post-vaccine neutralising antibody; SARS-S = Stimulated with SARS-CoV-2 peptide; CD4% = percent of CD4+ T cells; CD8% = percent of CD8+ T cells; CM = central memory; EM = effector memory; EMRA = Effector memory re-expressing CD45RA; WB = whole blood stimulation assay; CRP = C Reactive protein; LPS = lipopolysaccharide stimulated; CD3stim= PBMCs stimulated with anti-CD3.

| **Analyte** | **F-statistic** | **p-value** | **q-value** |
| --- | --- | --- | --- |
| **Neut Ab** | **14.75143623** | **0.000002510** | **0.000112931** |
| **CD4% SARS-S IL-2 IFNy** | **8.903148651** | **0.000280028** | **0.006300625** |
| **CD4% SARS-S TNF IL-2** | **8.350981712** | **0.000447823** | **0.006717338** |
| **CD4% SARS-S TNF IFNy** | **7.354140282** | **0.001057407** | **0.009634061** |
| **WB SARS-S IFNy** | **7.340041637** | **0.001070451** | **0.009634061** |
| CD4% SARS-S IL-2 | 4.408493519 | 0.014679182 | 0.110093862 |
| CD4% CM | 0.661622703 | 0.518302761 | 0.915459568 |
| CD4% EM | 0.43418476 | 0.649032549 | 0.915459568 |
| CD4% EMRA | 0.97393471 | 0.381218715 | 0.915459568 |
| CD4% CD3stim IFNy | 0.533860683 | 0.588032301 | 0.915459568 |
| CD4% CD3stim IL-2 IFNy | 1.042005539 | 0.356621314 | 0.915459568 |
| CD4% CD3stim TNF IFNy | 0.610551715 | 0.545103709 | 0.915459568 |
| CD4% CD3stim TNF IL-2 | 0.35597381 | 0.701392942 | 0.915459568 |
| CD4% SARS-S TNF | 0.340823352 | 0.712024108 | 0.915459568 |
| CD8% CM | 1.465167642 | 0.23605523 | 0.915459568 |
| CD8% EM | 0.35681048 | 0.700810586 | 0.915459568 |
| CD8% EMRA | 1.355161548 | 0.262695259 | 0.915459568 |
| CD8% CD3stim IFNy | 1.322524309 | 0.271174859 | 0.915459568 |
| CD8% CD3stim IL-2 | 1.122089267 | 0.329745445 | 0.915459568 |
| CD8% CD3stim TNF | 0.590034723 | 0.556264973 | 0.915459568 |
| CD8% CD3stim TNF IFNy | 1.095577955 | 0.338405755 | 0.915459568 |
| CD8% CD3stim TNF IL-2 | 0.942824781 | 0.393030177 | 0.915459568 |
| CD8% SARS-S IFNy | 0.99430573 | 0.37368131 | 0.915459568 |
| CD8% SARS-S IL-2 IFNy | 0.347255528 | 0.70749067 | 0.915459568 |
| CD8% SARS-S TNF | 0.695522726 | 0.501258821 | 0.915459568 |
| CD8% SARS-S TNF IFNy | 0.579966128 | 0.561827329 | 0.915459568 |
| CD8% SARS-S TNF IL-2 | 1.058355212 | 0.350958456 | 0.915459568 |
| CRP | 0.408156067 | 0.666000048 | 0.915459568 |
| WB LPS IFNy | 0.659529507 | 0.519374345 | 0.915459568 |
| WB LPS IL-6 | 0.411960125 | 0.663492304 | 0.915459568 |
| WB LPS IL-8 | 1.36215663 | 0.260913333 | 0.915459568 |
| WB SARS-S IL-6 | 1.501778722 | 0.227814238 | 0.915459568 |
| WB SARS-S IL-8 | 1.356493115 | 0.262355101 | 0.915459568 |
| WB SARS-S TNF | 0.866638422 | 0.423558472 | 0.915459568 |
| WB TNF | 0.485486239 | 0.616870022 | 0.915459568 |
| CD4% naive | 0.311279863 | 0.733230398 | 0.916537997 |
| CD4% CD3stim TNF | 0.217790619 | 0.804682057 | 0.928479296 |
| CD8% SARS-S IL-2 | 0.242988914 | 0.784751311 | 0.928479296 |
| WB IL6 | 0.237713635 | 0.788881718 | 0.928479296 |
| CD4% SARS-S IFNy | 0.177802429 | 0.837377229 | 0.942049383 |
| CD4% CD3stim IL-2 | 0.041483454 | 0.959382046 | 0.959400468 |
| CD8% naive | 0.051724799 | 0.949616066 | 0.959400468 |
| CD8% CD3stim IL-2 IFNy | 0.122665226 | 0.884695338 | 0.959400468 |
| WB LPS TNF | 0.079138882 | 0.923970584 | 0.959400468 |
| WB IL8 | 0.041464236 | 0.959400468 | 0.959400468 |

**Supplementary Table 2: Multivariate analysis of post-vaccine anti-S IgG/A/M antibody ratio and immune correlates after SARS-CoV-2 vaccination**

F-statistic (mean sum of squares regression / mean sum of squares error) represents the degree of relatedness of each immunological parameter to post-vaccine anti-S IgG/A/M antibody levels. p values and q values (false discovery rate) derived using the quadratic regression for general linear models with adjustment for the following covariates: age, sex, BMI category, vaccine types, inter-vaccine days, days post second vaccine, pre-vaccine SARS-CoV-2 sero-status, vitamin D allocation.

Neut Ab = post-vaccine neutralising antibody; SARS-S = Stimulated with SARS-CoV-2 peptide; CD4% = percent of CD4+ T cells; CD8% = percent of CD8+ T cells; CM = central memory; EM = effector memory; EMRA = Effector memory re-expressing CD45RA; WB = whole blood stimulation assay; CRP = C Reactive protein; LPS = lipopolysaccharide stimulated; CD3stim= PBMCs stimulated with anti-CD3.

| **Analyte** | **F-statistic** | **p-value** | **q-value** |
| --- | --- | --- | --- |
| **anti-S** | **18.82341003** | **1.37E-07** | **6.15E-06** |
| **CD8% SARS-S TNF** | **9.043599129** | **0.00025768** | **0.005797797** |
| WB LPS IFNg | 4.476495266 | 0.013927031 | 0.208905463 |
| CD4% SARS-S IL-2 IFNg | 4.155908108 | 0.018675077 | 0.210094621 |
| CD4% SARS-S TNF IFNg | 3.773694038 | 0.026559214 | 0.239032924 |
| CD4% SARS-S TNF IL-2 | 2.733723402 | 0.070201699 | 0.478347739 |
| CD8% CD3stim TNF | 2.672126532 | 0.074409648 | 0.478347739 |
| CD8% SARS-S IFNg | 2.120285988 | 0.125761624 | 0.707409137 |
| CD8% naive | 1.922573924 | 0.151997455 | 0.759987273 |
| WB LPS IL-6 | 1.616731644 | 0.204076375 | 0.865460951 |
| WB LPS IL-8 | 1.579493642 | 0.211557121 | 0.865460951 |
| CD8% CD3stim TNF IL-2 | 1.480389357 | 0.232861184 | 0.873229441 |
| CD8% CD3stim IL-2 | 1.285854697 | 0.281282137 | 0.9144346 |
| CD8% SARS-S TNF IFNg | 1.2453866 | 0.292584657 | 0.9144346 |
| CRP | 1.203369021 | 0.304811533 | 0.9144346 |
| CD4% EMRA | 0.43726778 | 0.647119287 | 0.979267193 |
| CD4% CD3stim IFNg | 0.671686947 | 0.513306603 | 0.979267193 |
| CD4% CD3stim IL-2 | 0.423572212 | 0.655960522 | 0.979267193 |
| CD4% CD3stim TNF | 0.556580186 | 0.575063077 | 0.979267193 |
| CD4% CD3stim TNF IFNg | 0.639137626 | 0.530049424 | 0.979267193 |
| CD4% CD3stim TNF IL-2 | 0.743589699 | 0.478209243 | 0.979267193 |
| CD4% SARS-S IFNg | 0.705029428 | 0.496715404 | 0.979267193 |
| CD8% EM | 0.514346242 | 0.59958208 | 0.979267193 |
| CD8% EMRA | 0.940470517 | 0.39412536 | 0.979267193 |
| CD8% CD3stim IL-2 IFNg | 0.792571187 | 0.455712813 | 0.979267193 |
| CD8% CD3stim TNF IFNg | 0.395296782 | 0.674606289 | 0.979267193 |
| CD8% SARS-S TNF IL-2 | 0.733572781 | 0.482947855 | 0.979267193 |
| WB LPS TNF | 0.879765749 | 0.418304014 | 0.979267193 |
| WB SARS-S IFNg | 0.863521218 | 0.425027598 | 0.979267193 |
| WB SARS-S IL-8 | 0.406051397 | 0.667451352 | 0.979267193 |
| WB SARS-S TNF | 0.497693449 | 0.609541115 | 0.979267193 |
| CD4% EM | 0.170627251 | 0.843399134 | 0.998762132 |
| CD4% naive | 0.224338278 | 0.799475929 | 0.998762132 |
| CD4% SARS-S IL-2 | 0.171836704 | 0.842383437 | 0.998762132 |
| CD4% SARS-S TNF | 0.274259329 | 0.76074756 | 0.998762132 |
| CD8% CD3stim IFNg | 0.256668478 | 0.774170844 | 0.998762132 |
| CD8% SARS-S IL-2 | 0.221605882 | 0.801652948 | 0.998762132 |
| WB SARS-S IL-6 | 0.262439609 | 0.769740529 | 0.998762132 |
| CD4% CM | 0.075825788 | 0.927034931 | 0.999529529 |
| CD4% CD3stim IL-2 IFNg | 0.03582282 | 0.96482453 | 0.999529529 |
| CD8% CM | 0.109211959 | 0.896655177 | 0.999529529 |
| CD8% SARS-S IL-2 IFNg | 0.045929633 | 0.955130819 | 0.999529529 |
| WB IL6 | 0.004542075 | 0.995468446 | 0.999529529 |
| WB IL8 | 0.063582912 | 0.938437068 | 0.999529529 |
| WB TNF | 0.000470584 | 0.999529529 | 0.999529529 |

**Supplementary Table 3: Multivariate analysis of post-vaccine neutralising antibody titre and immune correlates after SARS-CoV-2 vaccination**

F-statistic (mean sum of squares regression / mean sum of squares error) represents the degree of relatedness of each immunological parameter to post-vaccine neutralising antibody titre. p values and q values (false discovery rate) derived using the quadratic regression for general linear models with adjustment for the following covariates: age, sex, BMI category, vaccine types, inter-vaccine days, days post second vaccine, pre-vaccine SARS-CoV-2 sero-status, vitamin D allocation.

anti-S = anti-S IgG/A/M antibody ratio; SARS-S = Stimulated with SARS-CoV-2 peptide; CD4% = percent of CD4+ T cells; CD8% = percent of CD8+ T cells; CM = central memory; EM = effector memory; EMRA = Effector memory re-expressing CD45RA; WB = whole blood stimulation assay; CRP = C Reactive protein; LPS = lipopolysaccharide stimulated; CD3stim= PBMCs stimulated with anti-CD3.

| **Analyte** | **F-statistic** | **p-value** | **q-value** |
| --- | --- | --- | --- |
| **CD4% SARS-S IFNy** | **18.73680687** | **1.20E-07** | **5.52E-06** |
| **CD8% SARS-S IFNy** | **7.474424839** | **0.000939234** | **0.021602388** |
| **CD4% EM** | **5.325723648** | **0.006325274** | **0.096987535** |
| anti-S | 3.988625288 | 0.021516376 | 0.247438319 |
| CD4% EMRA | 3.007666111 | 0.053852537 | 0.495443344 |
| CD4% CM | 0.517386436 | 0.597647587 | 0.942721338 |
| CD4% naive | 1.409181714 | 0.249105527 | 0.942721338 |
| CD4% CD3stim IFNy | 0.995576322 | 0.373107074 | 0.942721338 |
| CD4% CD3stim IL-2 | 0.475562036 | 0.62292024 | 0.942721338 |
| CD4% CD3stim IL-2 IFNy | 0.072031185 | 0.930549634 | 0.942721338 |
| CD4% CD3stim TNF | 0.288744152 | 0.74982022 | 0.942721338 |
| CD4% CD3stim TNF IFNy | 0.865744114 | 0.423836746 | 0.942721338 |
| CD4% CD3stim TNF IL-2 | 0.747187436 | 0.476296963 | 0.942721338 |
| CD4% SARS-S IL-2 | 0.066743843 | 0.935476012 | 0.942721338 |
| CD4% SARS-S IL-2 IFNy | 0.525276363 | 0.59299855 | 0.942721338 |
| CD4% SARS-S TNF | 0.176050499 | 0.838832411 | 0.942721338 |
| CD4% SARS-S TNF IFNy | 0.590469599 | 0.555968161 | 0.942721338 |
| CD4% SARS-S TNF IL-2 | 0.954232514 | 0.388551695 | 0.942721338 |
| CD8% CM | 0.589098811 | 0.556721988 | 0.942721338 |
| CD8% EM | 0.442945182 | 0.643383771 | 0.942721338 |
| CD8% EMRA | 0.813568294 | 0.446156969 | 0.942721338 |
| CD8% naive | 0.30464676 | 0.738058985 | 0.942721338 |
| CD8% CD3stim IFNy | 0.974929035 | 0.380740336 | 0.942721338 |
| CD8% CD3stim IL-2 | 0.297078729 | 0.743632498 | 0.942721338 |
| CD8% CD3stim IL-2 IFNy | 0.14428176 | 0.865821908 | 0.942721338 |
| CD8% CD3stim TNF | 1.172135949 | 0.313881471 | 0.942721338 |
| CD8% CD3stim TNF IFNy | 0.974563479 | 0.380876907 | 0.942721338 |
| CD8% CD3stim TNF IL-2 | 0.07413473 | 0.928597062 | 0.942721338 |
| CD8% SARS-S IL-2 | 0.971634328 | 0.381973036 | 0.942721338 |
| CD8% SARS-S IL-2 IFNy | 0.217366144 | 0.805010724 | 0.942721338 |
| CD8% SARS-S TNF | 0.101282038 | 0.90376979 | 0.942721338 |
| CD8% SARS-S TNF IFNy | 0.41681692 | 0.660270179 | 0.942721338 |
| CD8% SARS-S TNF IL-2 | 0.132049665 | 0.876448511 | 0.942721338 |
| CRP | 0.059019007 | 0.942721338 | 0.942721338 |
| WB LPS IFNy | 1.56291151 | 0.214549643 | 0.942721338 |
| WB LPS IL-6 | 2.045507908 | 0.134637823 | 0.942721338 |
| WB LPS IL-8 | 1.355641603 | 0.262431513 | 0.942721338 |
| WB LPS TNF | 0.505159855 | 0.604925493 | 0.942721338 |
| Neut Ab | 0.202589706 | 0.816944248 | 0.942721338 |
| WB SARS-S IFNy | 0.259720981 | 0.771780272 | 0.942721338 |
| WB SARS-S IL-6 | 0.286217511 | 0.751706391 | 0.942721338 |
| WB SARS-S IL-8 | 0.204300418 | 0.815553483 | 0.942721338 |
| WB SARS-S TNF | 0.078542389 | 0.92451928 | 0.942721338 |
| WB IL6 | 0.355462164 | 0.701722691 | 0.942721338 |
| WB IL8 | 0.116096005 | 0.890508385 | 0.942721338 |
| WB TNF | 0.467428237 | 0.62796036 | 0.942721338 |

**Supplementary Table 4: Multivariate analysis of the effect of pre-vaccine anti-S IgG/A/M antibody ratio on immune correlates after SARS-CoV-2 vaccination**

F-statistic (mean sum of squares regression / mean sum of squares error) represents the degree of relatedness of each immunological parameter to pre-vaccine anti-S IgG/A/M antibody ratio. p values and q values (false discovery rate) derived using the quadratic regression for general linear models with adjustment for the following covariates: age, sex, BMI category, vaccine types, inter-vaccine days, days post second vaccine, pre-vaccine SARS-CoV-2 sero-status, vitamin D allocation.

Neut Ab = post-vaccine neutralising antibody; SARS-S = Stimulated with SARS-CoV-2 peptide; CD4% = percent of CD4+ T cells; CD8% = percent of CD8+ T cells; CM = central memory; EM = effector memory; EMRA = Effector memory re-expressing CD45RA; WB = whole blood stimulation assay; CRP = C Reactive protein; LPS = lipopolysaccharide stimulated; CD3stim= PBMCs stimulated with anti-CD3.

| **Analyte** | **t-statistic** | **p-value** | **q-value** |
| --- | --- | --- | --- |
| **anti-S** | **4.521405697** | **0.000015** | **0.000700526** |
| **Neut Ab** | **4.067364216** | **0.000088** | **0.002030986** |
| LPS TNF | 2.164852619 | 0.032502356 | 0.498369455 |
| CD4% CM | 1.731644273 | 0.08606595 | 0.728300214 |
| CD4% SARS-S IL-2 | -1.62280798 | 0.107417134 | 0.728300214 |
| CD8% EM | 1.680470109 | 0.095628286 | 0.728300214 |
| CD8% SARS-S IL-2 | -1.607076764 | 0.110828293 | 0.728300214 |
| CD4% naive | -1.469739676 | 0.144411041 | 0.743779567 |
| CD4% CD3stim TNF IL-2 | -1.408626676 | 0.16169121 | 0.743779567 |
| CD8% naive | -1.460416079 | 0.146950573 | 0.743779567 |
| CD4% CD3stim TNF | -1.348911285 | 0.180062418 | 0.752988293 |
| CD4% CD3stim IL-2 | -0.835657358 | 0.40511117 | 0.776463076 |
| CD4% CD3stim IL-2 IFNy | -0.902766705 | 0.368570093 | 0.776463076 |
| CD4% SARS-S IFNy | 0.856334627 | 0.393625003 | 0.776463076 |
| CD4% SARS-S TNF | -0.876020789 | 0.382877132 | 0.776463076 |
| CD4% SARS-S TNF IFNy | -0.878779888 | 0.381385464 | 0.776463076 |
| CD8% CM | 1.059227824 | 0.291754954 | 0.776463076 |
| CD8% CD3stim TNF | -1.173734069 | 0.242969483 | 0.776463076 |
| CD8% SARS-S TNF IL-2 | -1.008307815 | 0.315460952 | 0.776463076 |
| WB LPS IFNy | -0.968313694 | 0.33495569 | 0.776463076 |
| WB LPS IL-6 | 1.192560792 | 0.23553964 | 0.776463076 |
| WB LPS IL-8 | 1.033303618 | 0.303668241 | 0.776463076 |
| WB SARS-S IFNy | -0.891350746 | 0.374635055 | 0.776463076 |
| WB SARS-S TNF | -1.114505649 | 0.267427148 | 0.776463076 |
| CD4% SARS-S TNF IL-2 | -0.769994915 | 0.44290964 | 0.793774022 |
| WB IL6 | -0.760306478 | 0.448654882 | 0.793774022 |
| CD4% EM | 0.64762646 | 0.518540194 | 0.795094965 |
| CD8% CD3stim IL-2 IFNg | -0.671850979 | 0.50304986 | 0.795094965 |
| CD8% SARS-S IL-2 IFNg | -0.726016045 | 0.469330016 | 0.795094965 |
| WB IL8 | -0.701652944 | 0.484338226 | 0.795094965 |
| CD8% SARS-S TNF IFNy | -0.475702524 | 0.635204667 | 0.942561764 |
| CD8% CD3stim TNF IL-2 | -0.418027163 | 0.676720571 | 0.943307462 |
| CRP | -0.420758009 | 0.674730932 | 0.943307462 |
| CD4% CD3stim IFNy | -0.3179515 | 0.751108552 | 0.959749817 |
| CD4% CD3stim TNF IFNy | -0.355161577 | 0.7231306 | 0.959749817 |
| WB SARS-S IL-6 | -0.331206918 | 0.741101613 | 0.959749817 |
| CD8% CD3stim IL-2 | -0.259221524 | 0.795935969 | 0.963501437 |
| CD8% SARS-S TNF | 0.267031133 | 0.789931912 | 0.963501437 |
| CD4% SARS-S IL-2 IFNy | -0.190245017 | 0.849458485 | 0.967711782 |
| CD8% EMRA | 0.19088167 | 0.848960831 | 0.967711782 |
| CD8% SARS-S IFNy | -0.173555002 | 0.862525719 | 0.967711782 |
| CD4% EMRA | -0.032591127 | 0.974058143 | 0.985847417 |
| CD8% CD3stim IFNy | -0.03478156 | 0.97231529 | 0.985847417 |
| CD8% CD3stim TNF IFNy | -0.091835432 | 0.9269914 | 0.985847417 |
| WB SARS-S IL-8 | 0.017777862 | 0.985847417 | 0.985847417 |
| WB TNF | -0.046250455 | 0.963192267 | 0.985847417 |

**Supplementary Table 5: Multivariate analysis of effect of vaccine type on immune correlates after SARS-CoV-2 vaccination**

t-statistic (regression co-efficient / standard deviation) represents magnitude of difference between vaccine types; a positive t-statistic indicates a higher value and negative a lower value of immunological parameter in participants receiving BNT162b2 as compared to ChAdOx1-nCoV-19. p values and q values (false discovery rate) derived using the t-test for general linear models with adjustment for the following covariates: age, sex, BMI category, inter-vaccine days, days post second vaccine, pre-vaccine SARS-CoV-2 sero-status, vitamin D allocation.

anti-S = anti-S IgG/A/M antibody ratio; Neut Ab = post-vaccine neutralising antibody; SARS-S = Stimulated with SARS-CoV-2 peptide; CD4% = percent of CD4+ T cells; CD8% = percent of CD8+ T cells; CM = central memory; EM = effector memory; EMRA = Effector memory re-expressing CD45RA; WB = whole blood stimulation assay; CRP = C Reactive protein; LPS = lipopolysaccharide stimulated; CD3stim= PBMCs stimulated with anti-CD3.

| **Analyte** | **F-statistic** | **p-value** | **q-value** |
| --- | --- | --- | --- |
| CD4% SARS-S TNF IFNγ | 5.194693089 | 0.006959193 | 0.160061445 |
| CD8% SARS-S TNF | 5.339371681 | 0.006097158 | 0.160061445 |
| CD8% CD3stim TNF | 4.467357635 | 0.013594212 | 0.208444579 |
| CD4% CM | 3.067777395 | 0.050453844 | 0.580219205 |
| CD4% SARS-S IL-2 IFNγ | 2.734654188 | 0.069253577 | 0.607720777 |
| CD8% EM | 2.529609919 | 0.084235324 | 0.607720777 |
| CD8% CD3stim TNF IFNγ | 2.432103634 | 0.092479249 | 0.607720777 |
| CD4% EMRA | 1.382207036 | 0.255272258 | 0.780251089 |
| CD4% naive | 1.402085185 | 0.250368713 | 0.780251089 |
| CD4% CD3stim IL-2 | 1.199065924 | 0.305315644 | 0.780251089 |
| CD4% CD3stim TNF IL-2 | 1.210202098 | 0.302005272 | 0.780251089 |
| CD4% SARS-S IFNγ | 1.353111506 | 0.262626413 | 0.780251089 |
| CD4% SARS-S TNF IL-2 | 1.778921843 | 0.173556638 | 0.780251089 |
| CD8% CM | 1.241663337 | 0.292849029 | 0.780251089 |
| CD8% naive | 1.2514503 | 0.290058715 | 0.780251089 |
| CD8% CD3stim IFNγ | 1.570413232 | 0.212503503 | 0.780251089 |
| CD8% SARS-S IL-2 | 1.214286208 | 0.300800393 | 0.780251089 |
| WB TNF | 1.23462379 | 0.294872922 | 0.780251089 |
| CD4% EM | 0.943771005 | 0.392230136 | 0.828076871 |
| CD4% SARS-S IL-2 | 1.077064276 | 0.344095015 | 0.828076871 |
| CD8% SARS-S IFNγ | 0.954267919 | 0.388202363 | 0.828076871 |
| WB LPS IL-6 | 0.933950782 | 0.396036765 | 0.828076871 |
| CD4% CD3stim IL-2 IFNγ | 0.536748707 | 0.586142284 | 0.837857191 |
| CD4% CD3stim TNF | 0.485101759 | 0.616921982 | 0.837857191 |
| CD4% SARS-S TNF | 0.593892992 | 0.553902891 | 0.837857191 |
| CD8% CD3stim IL-2 | 0.706267893 | 0.495666831 | 0.837857191 |
| CD8% CD3stim IL-2 IFNγ | 0.828392446 | 0.439408789 | 0.837857191 |
| CD8% CD3stim TNF IL-2 | 0.664264917 | 0.516667324 | 0.837857191 |
| CD8% SARS-S IL-2 IFNγ | 0.490405381 | 0.613686826 | 0.837857191 |
| CD8% SARS-S TNF IL-2 | 0.527121723 | 0.591758709 | 0.837857191 |
| Neut Ab | 0.761065125 | 0.469569059 | 0.837857191 |
| WB IL6 | 0.507131457 | 0.603596738 | 0.837857191 |
| WB IL8 | 0.684206426 | 0.506586416 | 0.837857191 |
| anti-S | 0.481244534 | 0.61928575 | 0.837857191 |
| WB SARS-S IFNγ | 0.435780138 | 0.647851802 | 0.851462369 |
| WB LPS IFNγ | 0.386420488 | 0.680387106 | 0.869383525 |
| WB SARS-S IL-8 | 0.330656797 | 0.719150642 | 0.894079177 |
| CD4% CD3stim IFNγ | 0.264310747 | 0.768212454 | 0.90609674 |
| CRP | 0.28804189 | 0.750283883 | 0.90609674 |
| CD4% CD3stim TNF IFNγ | 0.203312948 | 0.816323329 | 0.915874954 |
| CD8% SARS-S TNF IFNγ | 0.226675957 | 0.79754388 | 0.915874954 |
| CD8% EMRA | 0.173164219 | 0.841224224 | 0.921340817 |
| WB SARS-S IL-6 | 0.122781418 | 0.88457585 | 0.924783844 |
| WB SARS-S TNF | 0.133890852 | 0.87482532 | 0.924783844 |
| WB LPS IL-8 | 0.088489637 | 0.915376525 | 0.935718225 |
| WB LPS TNF | 0.050919335 | 0.950377315 | 0.950377315 |

**Supplementary Table 6: Multivariate analysis of effect of the number of days between vaccine one and two on immune correlates after SARS-CoV-2 vaccination**

F-statistic (mean sum of squares regression / mean sum of squares error) represents the degree of relatedness of each immunological parameter to the number of inter-vaccine days. p values and q values (false discovery rate) derived using the quadratic regression for general linear models with adjustment for the following covariates: age, sex, BMI category, vaccine types, days post second vaccine, pre-vaccine SARS-CoV-2 sero-status, vitamin D allocation.

anti-S = anti-S IgG/A/M antibody ratio; Neut Ab = post-vaccine neutralising antibody; SARS-S = Stimulated with SARS-CoV-2 peptide; CD4% = percent of CD4+ T cells; CD8% = percent of CD8+ T cells; CM = central memory; EM = effector memory; EMRA = Effector memory re-expressing CD45RA; WB = whole blood stimulation assay; CRP = C Reactive protein; LPS = lipopolysaccharide stimulated; CD3stim= PBMCs stimulated with anti-CD3.

| **Analyte** | **t-statistic** | **p-value** | **q-value** |
| --- | --- | --- | --- |
| **WB SARS-S IL-6** | **5.816661358** | **6.81E-08** | **3.13E-06** |
| **WB SARS-S IL-8** | **5.316862106** | **6.16E-07** | **1.42E-05** |
| **WB SARS-S TNF** | **3.675478935** | **0.000379004** | **0.005811393** |
| Neut Ab | -2.54595637 | 0.012378562 | 0.142353462 |
| CD8% CD3stim IFNγ | -2.34131575 | 0.021141218 | 0.163344184 |
| CD8% CD3stim TNF IFNγ | -2.33826137 | 0.021305763 | 0.163344184 |
| CD4% EMRA | 2.202540159 | 0.029856457 | 0.196199577 |
| WB TNF | 2.07870698 | 0.040129141 | 0.230742562 |
| CD8% CD3stim IL-2 IFNγ | -2.0018158 | 0.047933894 | 0.244995456 |
| CD4% CM | -1.93632317 | 0.055568701 | 0.255616025 |
| CD8% CD3stim TNF | -1.7974751 | 0.075190405 | 0.260196162 |
| CD8% SARS-S TNF IL-2 | -1.77872109 | 0.078235703 | 0.260196162 |
| WB LPS IFNγ | 1.772967577 | 0.079190136 | 0.260196162 |
| anti-S | -1.85740244 | 0.066110782 | 0.260196162 |
| WB LPS IL-8 | -1.6853286 | 0.094952661 | 0.291188161 |
| CD4% SARS-S IFNγ | -1.51195717 | 0.133606936 | 0.362787116 |
| CD8% CD3stim IL-2 | -1.46333373 | 0.146421128 | 0.362787116 |
| CD8% SARS-S IL-2 | -1.4230845 | 0.157733529 | 0.362787116 |
| CD8% SARS-S TNF IFNγ | 1.441500068 | 0.152476937 | 0.362787116 |
| WB IL8 | 1.48661387 | 0.140171558 | 0.362787116 |
| CD8% SARS-S TNF | 1.257017016 | 0.211590477 | 0.443176692 |
| WB LPS TNF | -1.25600946 | 0.21195407 | 0.443176692 |
| CD8% CD3stim TNF IL-2 | -1.09772456 | 0.274884366 | 0.512657872 |
| CD8% SARS-S IL-2 IFNγ | -1.08917522 | 0.278618409 | 0.512657872 |
| WB LPS IL-6 | 1.111456633 | 0.268959521 | 0.512657872 |
| CD4% SARS-S IL-2 | -0.82717258 | 0.410051507 | 0.723345024 |
| WB IL6 | 0.801700234 | 0.424572079 | 0.723345024 |
| CD4% EM | 0.737711966 | 0.46236686 | 0.759602699 |
| CD4% SARS-S TNF | 0.602979541 | 0.547847707 | 0.868999812 |
| CD4% CD3stim IL-2 | -0.43791673 | 0.662363631 | 0.897725744 |
| CD4% CD3stim TNF | 0.409460247 | 0.683052197 | 0.897725744 |
| CD4% CD3stim TNF IL-2 | -0.42724279 | 0.670094318 | 0.897725744 |
| CD8% CM | -0.41620389 | 0.678126783 | 0.897725744 |
| CD8% EM | -0.50168902 | 0.616956977 | 0.897725744 |
| CD8% SARS-S IFNγ | 0.467321068 | 0.641257333 | 0.897725744 |
| CD4% CD3stim IL-2 IFNγ | -0.35103887 | 0.726276186 | 0.899126403 |
| CD4% SARS-S TNF IL-2 | -0.30959654 | 0.757493253 | 0.899126403 |
| CD8% naive | 0.303260773 | 0.76230282 | 0.899126403 |
| WB SARS-S IFNγ | 0.338321298 | 0.735809726 | 0.899126403 |
| CRP | -0.25144184 | 0.801973502 | 0.922269527 |
| CD4% naive | 0.119311832 | 0.905260709 | 0.925377614 |
| CD4% CD3stim IFNγ | -0.15342098 | 0.878366475 | 0.925377614 |
| CD4% CD3stim TNF IFNγ | -0.12735347 | 0.89890894 | 0.925377614 |
| CD4% SARS-S TNF IFNγ | 0.166349515 | 0.868208013 | 0.925377614 |
| CD8% EMRA | -0.13402428 | 0.893644905 | 0.925377614 |
| CD4% SARS-S IL-2 IFNγ | -0.09183755 | 0.92700549 | 0.92700549 |

**Supplementary Table 7: Multivariate analysis of correlation between number of days after the second vaccine on immune correlates after SARS-CoV-2 vaccination**

t-statistic (regression co-efficient / standard deviation) represents magnitude of relationship between each immunological parameter to the number of days after the second vaccine and blood sampling. p values and q values (false discovery rate) derived using linear regression for general linear models with adjustment for the following covariates: age, sex, BMI category, vaccine types, pre-vaccine SARS-CoV-2 sero-status, vitamin D allocation.

anti-S = anti-S IgG/A/M antibody ratio; Neut Ab = post-vaccine neutralising antibody; SARS-S = Stimulated with SARS-CoV-2 peptide; CD4% = percent of CD4+ T cells; CD8% = percent of CD8+ T cells; CM = central memory; EM = effector memory; EMRA = Effector memory re-expressing CD45RA; WB = whole blood stimulation assay; CRP = C Reactive protein; LPS = lipopolysaccharide stimulated; CD3stim= PBMCs stimulated with anti-CD3.

| **Analyte** | **t-statistic** | **p-value** | **q-value** |
| --- | --- | --- | --- |
| **CD8% naive** | **-6.228470802** | **1.04E-08** | **4.78E-07** |
| **CD8% EM** | **2.97858429** | **0.003611396** | **0.042298032** |
| **CD8% EMRA** | **2.972481966** | **0.00367809** | **0.042298032** |
| **anti-S** | **-3.139087915** | **0.002211252** | **0.042298032** |
| **CD4% SARS-S IL-2 IFNγ** | **-2.549304485** | **0.012267475** | **0.09590231** |
| **CD4% SARS-S TNF IL-2** | **-2.542059422** | **0.012508997** | **0.09590231** |
| CD4% CD3stim IL-2 | -2.006546497 | 0.047418833 | 0.311609471 |
| CD4% CD3stim TNF IL-2 | -1.905206323 | 0.05954217 | 0.342367477 |
| CD8% CM | 1.791730642 | 0.076112588 | 0.389019895 |
| CD4% EM | 1.181297064 | 0.240205144 | 0.587892344 |
| CD4% naive | -1.288634181 | 0.20041226 | 0.587892344 |
| CD4% SARS-S TNF IFNγ | -1.348785639 | 0.180363843 | 0.587892344 |
| CD8% CD3stim IL-2 IFNγ | -1.17469573 | 0.242825099 | 0.587892344 |
| CD8% CD3stim TNF | 1.303633928 | 0.195264679 | 0.587892344 |
| CD8% CD3stim TNF IL-2 | -1.454822659 | 0.148759184 | 0.587892344 |
| CD8% SARS-S TNF IFNγ | 1.19952631 | 0.23307542 | 0.587892344 |
| Neut Ab | -1.24086082 | 0.217476018 | 0.587892344 |
| WB SARS-S IL-8 | -1.446853518 | 0.15097449 | 0.587892344 |
| WB SARS-S TNF | -1.356435537 | 0.177926461 | 0.587892344 |
| CD4% SARS-S IFNγ | 1.004557252 | 0.317464248 | 0.7014113 |
| CD8% CD3stim IL-2 | -0.976980507 | 0.330867861 | 0.7014113 |
| CD8% CD3stim TNF IFNγ | 0.937478244 | 0.35070565 | 0.7014113 |
| WB LPS TNF | 0.962249219 | 0.338178038 | 0.7014113 |
| CD4% CM | 0.872027755 | 0.385221441 | 0.738341096 |
| WB LPS IL-8 | -0.833280206 | 0.406614865 | 0.748171351 |
| CD8% SARS-S TNF | 0.782918513 | 0.435471371 | 0.770449349 |
| CD8% SARS-S IFNγ | 0.747615278 | 0.456395709 | 0.77756306 |
| CD8% CD3stim IFNγ | 0.691912115 | 0.490549318 | 0.778112711 |
| WB SARS-S IL-6 | -0.710655749 | 0.478903673 | 0.778112711 |
| CD8% SARS-S IL-2 IFNγ | -0.649149895 | 0.517686388 | 0.793785794 |
| WB IL8 | -0.590743661 | 0.555986234 | 0.825011832 |
| CRP | 0.549275875 | 0.584005033 | 0.839507235 |
| CD4% CD3stim TNF | -0.437128425 | 0.662933331 | 0.924088886 |
| WB LPS IL-6 | 0.359537393 | 0.719929186 | 0.9268921 |
| SARS-S IFNγ | -0.352218807 | 0.725393817 | 0.9268921 |
| WB IL6 | -0.405275166 | 0.686115808 | 0.9268921 |
| CD4% EMRA | 0.277177006 | 0.782199713 | 0.938712658 |
| CD8% SARS-S IL-2 | 0.259373099 | 0.79586508 | 0.938712658 |
| WB TNF | 0.304923832 | 0.761039459 | 0.938712658 |
| CD4% CD3stim IL-2 IFNγ | -0.153126284 | 0.878598276 | 0.962274302 |
| CD8% SARS-S TNF IL-2 | -0.156635746 | 0.875838541 | 0.962274302 |
| WB LPS IFNγ | 0.200240687 | 0.841687181 | 0.962274302 |
| CD4% CD3stim IFNγ | -0.050171085 | 0.960083218 | 0.980254766 |
| CD4% CD3stim TNF IFNγ | -0.047316428 | 0.962352658 | 0.980254766 |
| CD4% SARS-S IL-2 | 0.024809688 | 0.980254766 | 0.980254766 |
| CD4% SARS-S TNF | -0.032009501 | 0.97452642 | 0.980254766 |

**Supplementary Table 8: Multivariate analysis of correlation between age and immune correlates after SARS-CoV-2 vaccination**

t-statistic (regression co-efficient / standard deviation) represents magnitude of relationship between each immunological parameter to year of age. p values and q values (false discovery rate) derived using linear regression for general linear models with adjustment for the following covariates: sex, BMI category, vaccine types, inter-vaccine days, days post second vaccine, pre-vaccine SARS-CoV-2 sero-status, vitamin D allocation.

anti-S = anti-S IgG/A/M antibody ratio; Neut Ab = post-vaccine neutralising antibody; SARS-S = Stimulated with SARS-CoV-2 peptide; CD4% = percent of CD4+ T cells; CD8% = percent of CD8+ T cells; CM = central memory; EM = effector memory; EMRA = Effector memory re-expressing CD45RA; WB = whole blood stimulation assay; CRP = C Reactive protein; LPS = lipopolysaccharide stimulated; CD3stim= PBMCs stimulated with anti-CD3.

| **Analyte** | **F-statistic** | **p-value** | **q-value** |
| --- | --- | --- | --- |
| **CD4% SARS-S TNF** | **13.34089279** | **2.01E-07** | **9.24E-06** |
| **CD8% SARS-S TNF** | **9.893043518** | **8.65E-06** | **0.000198858** |
| CD4% SARS-S IL-2 | 3.781968117 | 0.012769521 | 0.155897819 |
| CRP | 3.734163523 | 0.013556332 | 0.155897819 |
| CD4% EM | 2.265696049 | 0.085278286 | 0.435866793 |
| CD8% CM | 2.331936359 | 0.07851803 | 0.435866793 |
| CD8% EMRA | 2.416109085 | 0.070686109 | 0.435866793 |
| CD8% CD3stim IL-2 | 2.436052084 | 0.068946709 | 0.435866793 |
| CD8% SARS-S IL-2 | 2.339568377 | 0.077773932 | 0.435866793 |
| CD4% CM | 1.923565388 | 0.130395817 | 0.487805718 |
| CD4% SARS-S TNF IL-2 | 1.736675382 | 0.164126928 | 0.487805718 |
| CD8% CD3stim IFNγ | 1.745974183 | 0.162265564 | 0.487805718 |
| CD8% CD3stim TNF IL-2 | 2.028518438 | 0.114515798 | 0.487805718 |
| CD8% SARS-S IL-2 IFNγ | 1.898503065 | 0.134494113 | 0.487805718 |
| WB LPS IFNγ | 1.84949255 | 0.142873201 | 0.487805718 |
| WB LPS IL-6 | 1.660010099 | 0.180276026 | 0.487805718 |
| WB LPS TNF | 1.694372058 | 0.172857538 | 0.487805718 |
| CD4% EMRA | 1.499236465 | 0.219217646 | 0.504200585 |
| CD8% CD3stim IL-2 IFNγ | 1.570018411 | 0.20117689 | 0.504200585 |
| CD8% CD3stim TNF IFNγ | 1.501374364 | 0.218650964 | 0.504200585 |
| CD4% naive | 1.317798018 | 0.272678919 | 0.507418637 |
| CD4% CD3stim IFNγ | 1.308351755 | 0.275770999 | 0.507418637 |
| CD4% CD3stim IL-2 IFNγ | 1.437575936 | 0.236171489 | 0.507418637 |
| CD4% CD3stim TNF IFNγ | 1.368216634 | 0.256712754 | 0.507418637 |
| CD8% SARS-S TNF IL-2 | 1.347492218 | 0.263167276 | 0.507418637 |
| CD8% CD3stim TNF | 1.065312862 | 0.367237763 | 0.64972835 |
| CD4% SARS-S IL-2 IFNγ | 1.029613495 | 0.382752401 | 0.652096684 |
| anti-S | 0.956532121 | 0.416307417 | 0.683933614 |
| WB SARS-S TNF | 0.837654352 | 0.476235709 | 0.755408367 |
| CD8% naive | 0.733924806 | 0.534101323 | 0.792537448 |
| WB TNF | 0.750576198 | 0.52445981 | 0.792537448 |
| CD4% CD3stim TNF | 0.68251729 | 0.564710995 | 0.799612798 |
| CD4% SARS-S IFNγ | 0.623096943 | 0.601649702 | 0.799612798 |
| CD8% EM | 0.61251086 | 0.608401042 | 0.799612798 |
| WB SARS-S IL-8 | 0.62098521 | 0.602992433 | 0.799612798 |
| CD8% SARS-S TNF IFNγ | 0.504633486 | 0.679932034 | 0.845320907 |
| WB LPS IL-8 | 0.523700774 | 0.666947847 | 0.845320907 |
| CD4% SARS-S TNF IFNγ | 0.380030721 | 0.767595065 | 0.882734324 |
| CD8% SARS-S IFNγ | 0.388651192 | 0.76140946 | 0.882734324 |
| WB IL6 | 0.424523562 | 0.735822283 | 0.882734324 |
| CD4% CD3stim IL-2 | 0.292669058 | 0.830610472 | 0.931904432 |
| CD4% CD3stim TNF IL-2 | 0.183677852 | 0.907277768 | 0.93718602 |
| Neut Ab | 0.144979641 | 0.932683726 | 0.93718602 |
| WB SARS-S IFNγ | 0.210271448 | 0.889085492 | 0.93718602 |
| WB SARS-S IL-6 | 0.242032945 | 0.86682437 | 0.93718602 |
| WB IL8 | 0.137849733 | 0.93718602 | 0.93718602 |

**Supplementary Table 9: Multivariate analysis of association between BMI category and immune correlates after SARS-CoV-2 vaccination**

F-statistic (mean sum of squares regression / mean sum of squares error) represents the degree of relatedness of each immunological parameter to BMI category. p values and q values (false discovery rate) derived using ANOVA for general linear models with adjustment for the following covariates: age, sex, vaccine types, inter-vaccine days, days post second vaccine, pre-vaccine SARS-CoV-2 sero-status, vitamin D allocation.

anti-S = anti-S IgG/A/M antibody ratio; Neut Ab = post-vaccine neutralising antibody; SARS-S = Stimulated with SARS-CoV-2 peptide; CD4% = percent of CD4+ T cells; CD8% = percent of CD8+ T cells; CM = central memory; EM = effector memory; EMRA = Effector memory re-expressing CD45RA; WB = whole blood stimulation assay; CRP = C Reactiv e protein; LPS = lipopolysaccharide stimulated; CD3stim= PBMCs stimulated with anti-CD3.

| **Analyte** | **t-statistic** | **p-value** | **q-value** |
| --- | --- | --- | --- |
| **CD8% EM** | **4.012705803** | **0.000108231** | **0.002489318** |
| **WB LPS IL-6** | **4.158295155** | **6.27E-05** | **0.002489318** |
| **CD8% naive** | **-3.182844162** | **0.001883947** | **0.02888719** |
| **CD4% CM** | **3.01518178** | **0.003172075** | **0.036478867** |
| **WB LPS IL-8** | **2.913638115** | **0.004307056** | **0.039624915** |
| **CD4% EM** | **2.567760468** | **0.011541032** | **0.079635906** |
| **CD4% naive** | **-2.549779177** | **0.012118507** | **0.079635906** |
| CD8% CM | 2.294992447 | 0.023579684 | 0.135583181 |
| WB LPS TNF | 1.751663685 | 0.082544731 | 0.42189529 |
| CD4% EMRA | -1.522587419 | 0.130655571 | 0.441795653 |
| CD4% SARS-S IFNγ | -1.592615604 | 0.114039998 | 0.441795653 |
| CD4% SARS-S TNF IFNγ | 1.447651625 | 0.150483264 | 0.441795653 |
| WB SARS-S IL-6 | 1.616152287 | 0.108849931 | 0.441795653 |
| WB SARS-S IL-8 | 1.489691854 | 0.139091161 | 0.441795653 |
| WB SARS-S TNF | 1.490472198 | 0.138886246 | 0.441795653 |
| anti-S | 1.436341166 | 0.153668053 | 0.441795653 |
| CD8% SARS-S IL-2 | -1.355089664 | 0.178091714 | 0.481895225 |
| CD4% SARS-S IL-2 IFNγ | 1.222740889 | 0.223971666 | 0.542810819 |
| CD8% CD3stim TNF IFNγ | 1.22212255 | 0.224204469 | 0.542810819 |
| CD8% CD3stim IFNγ | 1.164691687 | 0.246596752 | 0.567172529 |
| CD4% SARS-S IL-2 | -1.080163479 | 0.282369432 | 0.618523518 |
| CD8% CD3stim TNF | 1.042273998 | 0.299509431 | 0.626246992 |
| CD4% CD3stim IFNγ | 0.588249743 | 0.557538563 | 0.802987041 |
| CD4% CD3stim IL-2 | -0.55022037 | 0.583254232 | 0.802987041 |
| CD4% CD3stim TNF IFNγ | 0.755102396 | 0.451758516 | 0.802987041 |
| CD4% SARS-S TNF | -0.593918622 | 0.553753652 | 0.802987041 |
| CD4% SARS-S TNF IL-2 | 0.820012748 | 0.413935175 | 0.802987041 |
| CD8% SARS-S TNF | -0.560191274 | 0.576457882 | 0.802987041 |
| CD8% SARS-S TNF IFNγ | 0.664710104 | 0.507590186 | 0.802987041 |
| Neut Ab | 0.739851773 | 0.460924267 | 0.802987041 |
| WB SARS-S IFNγ | -0.604854286 | 0.546488507 | 0.802987041 |
| WB IL6 | 0.535274148 | 0.593512161 | 0.802987041 |
| WB IL8 | 0.612185717 | 0.541644781 | 0.802987041 |
| WB TNF | 0.751627922 | 0.45383747 | 0.802987041 |
| WB LPS IFNγ | 0.463643044 | 0.643795355 | 0.846131038 |
| CD4% CD3stim TNF IL-2 | -0.409734994 | 0.682776054 | 0.847087595 |
| CD8% EMRA | 0.397966474 | 0.691405752 | 0.847087595 |
| CD8% CD3stim IL-2 | -0.357531309 | 0.721360997 | 0.847087595 |
| CD8% CD3stim TNF IL-2 | -0.317172229 | 0.751698191 | 0.847087595 |
| CD8% SARS-S IFNγ | 0.367500424 | 0.713933175 | 0.847087595 |
| CRP | 0.312795132 | 0.755012857 | 0.847087595 |
| CD8% SARS-S IL-2 IFNγ | -0.281562924 | 0.77879364 | 0.852964463 |
| CD8% CD3stim IL-2 IFNγ | -0.245385736 | 0.806602876 | 0.862877496 |
| CD4% CD3stim IL-2 IFNγ | 0.164409906 | 0.869702196 | 0.889028912 |
| CD8% SARS-S TNF IL-2 | 0.181521699 | 0.856283308 | 0.889028912 |
| CD4% CD3stim TNF | 0.032635368 | 0.97402294 | 0.97402294 |

**Supplementary Table 10: Multivariate analysis of association between Sex and immune correlates after SARS-CoV-2 vaccination**

t-statistic (regression co-efficient / standard deviation) represents magnitude of difference between sexes; a positive t-statistic indicates a higher value and negative a lower value of immunological parameter in males participants compared to female. p values and q values (false discovery rate) derived using the t-test for general linear models with adjustment for the following covariates: age, BMI category, vaccine types, inter-vaccine days, days post second vaccine, pre-vaccine SARS-CoV-2 sero-status, vitamin D allocation.

anti-S = anti-S IgG/A/M antibody ratio; Neut Ab = post-vaccine neutralising antibody; SARS-S = Stimulated with SARS-CoV-2 peptide; CD4% = percent of CD4+ T cells; CD8% = percent of CD8+ T cells; CM = central memory; EM = effector memory; EMRA = Effector memory re-expressing CD45RA; WB = whole blood stimulation assay; CRP = C Reactive protein; LPS = lipopolysaccharide stimulated; CD3stim= PBMCs stimulated with anti-CD3.
